# A genome-wide association study of mammographic texture variation

**DOI:** 10.1101/2022.07.25.22278024

**Authors:** Yuxi Liu, Hongjie Chen, John Heine, Sara Lindstroem, Constance Turman, Erica T. Warner, Stacey J. Winham, Celine M. Vachon, Rulla M. Tamimi, Peter Kraft, Xia Jiang

**Author notes:** Corresponding authors: Peter Kraft, PhD. Program in Genetic Epidemiology and Statistical Genetics, Harvard T.H. Chan School of Public Health, 655 Huntington Avenue, Building 2-249A, Boston, MA, 02115., Xia Jiang, PhD. Department of Clinical Neuroscience, Center for Molecular Medicine, Karolinska Institutet, Visionsgatan 18, Solna 171 77, Stockholm, Sweden.

## Abstract

**Background:** Breast parenchymal texture features, including gray scale variation (V), capture the patterns of texture variation on a mammogram and are associated with breast cancer risk, independent of mammographic density (MD). However, our knowledge on the genetic basis of these texture features is limited.

**Methods:** We conducted a genome-wide association study of V in 7,040 European-ancestry women. Four V assessments representing different amounts of breast edge erosion and image resolutions were generated from digitized film mammograms. We used linear regression to test the single-nucleotide polymorphism (SNP)-phenotype associations adjusting for age, body mass index (BMI), MD phenotypes, and the top four genetic principal components. Multivariate phenotype association tests combining all four V assessments were performed. We further calculated genetic correlations and performed SNP-set tests of V with MD, breast cancer risk, and other breast cancer risk factors.

**Results:** We identified three genome-wide significant loci associated with V: rs138141444 (6q24.1) in *ECT2L*, rs79670367 (8q24.22) in *LINC01591*, and rs113174754 (12q22) near *PGAM1P5*. 6q24.1 and 8q24.22 have not previously been associated with MD phenotypes or breast cancer risk, whilst 12q22 is a known locus for both MD and breast cancer risk. Among known MD and breast cancer risk SNPs, we identified four variants that were associated with V at the Bonferroni-corrected thresholds accounting for the number of SNPs tested: rs335189 (5q23.2) in *PRDM6*, rs13256025 (8p21.2) in *EBF2*, rs11836164 (12p12.1) near *SSPN*, and rs17817449 (16q12.2) in *FTO*. We observed significant genetic correlations between V and mammographic dense area (r_g_ = 0.79, *P* = 5.91 × 10^−5^), percent density (r_g_ = 0.73, *P* = 1.00 × 10^−4^), and adult BMI (r_g_ = -0.36, *P* = 3.88 × 10^−7^). Additional significant relationships were observed for nondense area (z = -4.14, *P* = 3.42 × 10^−5^), estrogen receptor-positive breast cancer (z = 3.41, *P* = 6.41 × 10^−4^), and childhood body fatness (z = -4.91, *P* = 9.05 × 10^−7^) from the SNP-set tests.

**Conclusions:** These findings provide new insights into the genetic basis of mammographic texture variation and their associations with MD, breast cancer risk, and other breast cancer risk factors.

## Background

Mammographic density (MD) phenotypes reflect the amount of dense or nondense tissue on a mammogram and are well-established risk factors for breast cancer [1-3]. MD phenotypes are highly heritable with h^2^ = 60-70% from twin studies [4, 5]. Genome-wide association studies (GWAS) have identified 55 loci that are associated with MD phenotypes [6-8], including 32 loci for dense area (DA), which reflects the amount of fibroglandular tissue in the breast, 18 loci for nondense area (NDA), which reflects the amount of fatty tissue in the breast, and 24 loci for percent density (PD), defined as the percentage of area on a mammogram that is occupied by dense tissue [9].

Yet, MD is a global metric that ignores local patterns of variability in breast density [10]. Women with the same level of PD may have substantial heterogeneity in the structural patterns of breast parenchyma, which are assessed as texture features. Compared to MD phenotypes, breast parenchymal texture features are more refined and localized, and are fully automated measures of the variation in parenchymal patterns on a mammogram [11]. Growing evidence suggest that texture features are independent breast cancer risk factors [12-16]. Heine et al. developed a summary measure of texture features called V, which captures the gray scale variation on a mammogram [12]. Recent studies have shown that a higher value of V, reflecting greater texture variation, is associated with an increased risk of breast cancer, independent of MD [12, 16]. Understanding the mechanisms underlying texture variation and breast cancer risk, especially the role of genetic variants, would provide additional insights into the development of breast cancer. However, to date, no GWAS has been conducted on breast parenchymal texture features.

In the present study, we performed a GWAS of mammographic texture variation, including four different assessments of V, within the Nurses’ Health Studies and Mayo Mammography Health Study cohorts. We also leveraged summary statistics of breast cancer risk and MD phenotypes from previous GWAS to identify shared susceptibility loci for V, MD, and breast cancer risk. We further assessed the genetic relationships of V with MD phenotypes, breast cancer risk, and other breast cancer risk factors by estimating genetic correlations and performing single-nucleotide polymorphism (SNP)-set tests.

## Methods

### Study population

The Nurses’ Health Study (NHS) is a prospective cohort study established in 1976. A total number of 121,700 female registered nurses aged 30 to 55 residing in 11 states within the United States completed an initial questionnaire at that time. NHSII was established in 1989 when 116,671 female registered nurses aged 25 to 42 residing in 14 states completed an initial questionnaire. Blood samples were collected from 32,826 women in NHS cohort from 1989 to 1990 and 29,611 women in NHSII cohort from 1996 to 1999, which form the blood subcohorts. Women in each cohort have been followed by self-administered questionnaires to collect updated exposure and newly diagnosed disease information every two years.

The Mayo Mammography Health Study (MMHS) is a prospective cohort study of 19,924 women who had a screening mammogram from 2003 to 2006 at the Mayo Clinic in Rochester, MN and agreed to participate in the study. Participants were at least 35 years old, residents of Minnesota, Iowa, or Wisconsin, and had no personal history of breast cancer. Participants completed a baseline questionnaire and provided consent to access any residual blood samples from clinical tests over the time period. Breast cancer diagnostic information were obtained through linkage to state-wide cancer registry data and mailed questionnaires.

### Mammogram collection and processing

The mammogram collection and processing procedure in NHS and NHSII is described elsewhere [16, 17] and is briefly summarized here. Pre-diagnostic screening mammograms were collected within NHS and NHSII breast cancer case-control studies nested in the blood subcohorts [18]. A total number of 6,258 film mammograms obtained close to the blood draw date were initially collected. The study protocol was approved by the institutional review boards of the Brigham and Women’s Hospital and Harvard T.H. Chan School of Public Health. Film mammogram craniocaudal views of both breasts were digitized using a Lumysis 85 laser film scanner or a VIDAR CAD PRO Advantage scanner (VIDAR Systems Corporation, Herndon, VA, USA). Digitized images were grouped based on resolution (mean resolution = 171μm, 232μm, 300μm, and images with isolated resolutions). Here, we evaluated the groups of images with average resolutions of 171μm (high resolution) and 300μm (low resolution). Images with isolated resolutions were down-sampled to 300μm and added to the low resolution group. All 171μm images were further adjusted to 300μm to form a larger dataset of low-resolution images.

Details of mammogram acquisition, retrieval, and digitization for MMHS are described elsewhere [12, 19]. Briefly, women in MMHS who agreed to participate provided written informed consent to access their mammograms. A total number of 19,924 women were followed up for incident cancer events. We used a case-cohort design with a random sample of 2,300 women from the entire MMHS cohort as the subcohort. We collected film mammograms from 1,194 breast cancer cases identified through August 2019, excluding women who were diagnosed within 60 days of the enrollment mammogram and women with a digital mammogram. We further collected mammograms from 2,167 control women from the subcohort. The study protocol was approved by the Mayo Clinic institutional review board. Film mammograms of both craniocaudal views were digitized on the Array 2905 laser digitizer (Array Corporation, Roden, The Netherlands) with 50μm (limiting) pixel spacing and further downsampled to 200μm. Both the original 50μm images (high resolution) and the down-sampled 200μm images (low resolution) were used for calculation of V.

### Assessment of V

V is an automated measure of the gray scale variation on a mammogram. The algorithm for generating V has been described previously by Heine et al. [12, 20, 21]. Briefly, there are three main steps: segmentation, erosion, and calculation of variation. First, the breast is segmented from the background. Then, the segmented breast area is eroded by 25% or 35% along a radial direction to retain the regions where the breast was in contact with the compression paddle. Finally, the V is calculated as the standard deviation of the pixel values within the eroded breast region. Normalization processes, including spatial normalization, feature distribution normalization, and resolution estimation, were applied to the images before calculation of V to account for resolution and intensity scale differences [17].

We generated four assessments of V with different proportions of erosion and image resolutions: V with 35% erosion and low resolution (V65L), V with 25% erosion and low resolution (V75L), V with 35% erosion and high resolution (V65H), and V with 25% erosion and high resolution (V75H). These four V assessments were highly correlated with each other (Fig. 1a). We used V65L as our primary univariate outcome, as it had the largest sample size.

**Fig. 1.**
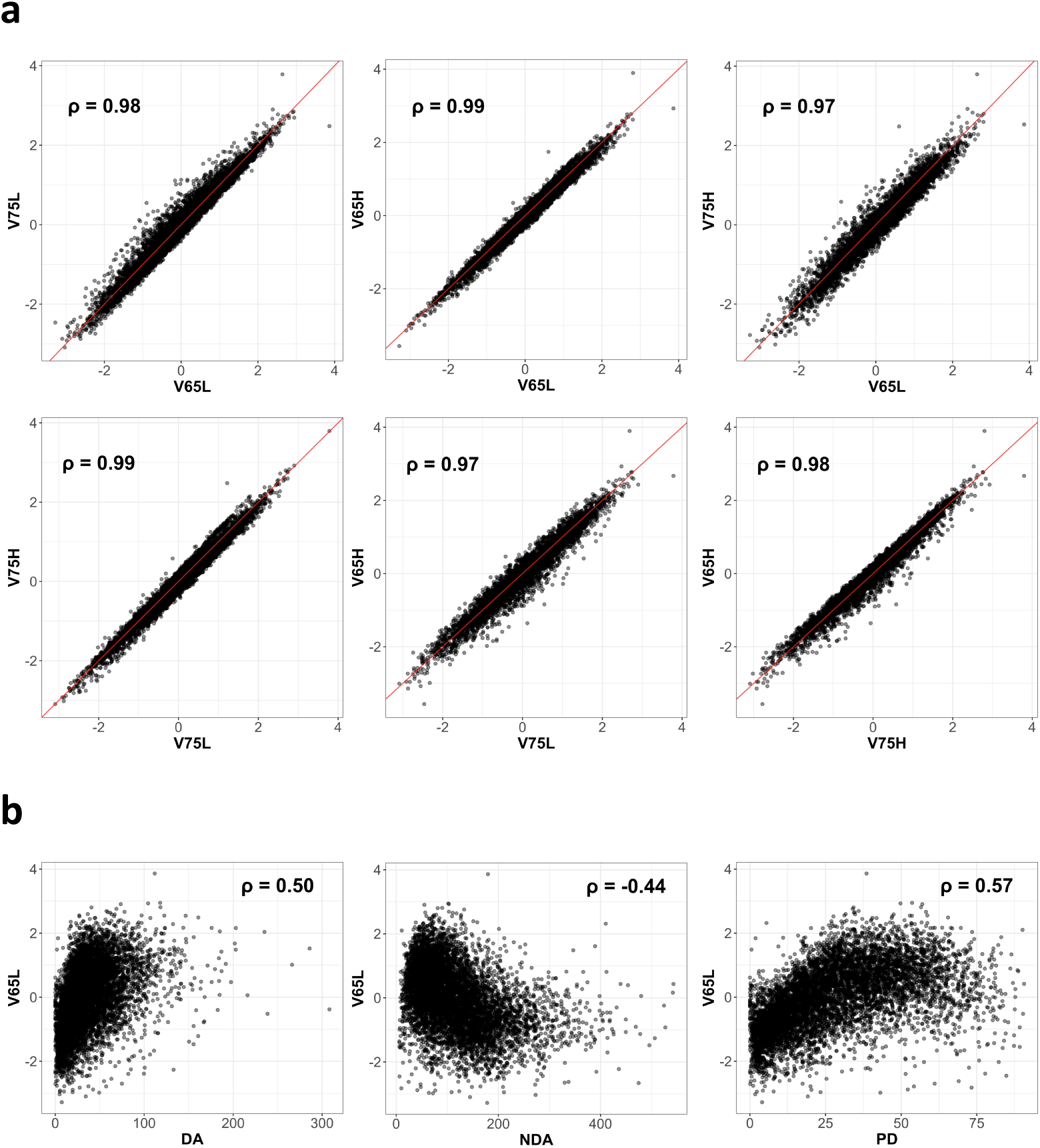
Scatter plots of four V assessments and V65L by three mammographic density phenotypes. **a** = scatter plots of four V assessments; **b** = scatter plots of V65L by dense area (DA), nondense area (NDA), and percent density (PD). Spearman correlation between the two measures is shown on the upper left or right corner of each plot. Red lines on the plots are the diagonal lines.

### MD phenotypes and other covariates

MD phenotypes were assessed from digitized film mammograms using Cumulus [22], a semi-automated software, by a single trained reader [12, 23]. DA and NDA were generated for each mammogram; PD was calculated as DA divided by the total breast area. DA, NDA, and PD measures in the left and right breasts were averaged. Fig. 1b shows the scatter plots and correlations of V65L and the three MD phenotypes. Body mass index (BMI) was measured at mammogram collection for all participants. Women were considered as breast cancer cases if they were diagnosed with breast cancer after blood or mammogram collection but before June 1, 2004 (NHS), June 1, 2007 (NHSII), or August 2019 (MMHS). Age at mammogram collection was also retrieved.

### Genotyping, quality control, and imputation

The full genotyping and quality control pipeline for NHS and NHSII is described elsewhere [24]. In the present study, we used genotype data from four platforms: ALymetrix 6.0, Illumina HumanHap, Illumina OmniExpress, and Illumina OncoArray. Variants with call rate < 95% or Hardy-Weinberg equilibrium *P* < 1 × 10^−6^ were excluded. European ancestry principal component (PC) outliers or samples with call rate < 90%, gender discordance, or extreme heterozygosity were excluded.

The full genotyping and quality control pipeline for MMHS is also described elsewhere [25]. Here, we used genotype data from iCOGS and OncoArray platforms. Variants with a call rate < 95% or not in Hardy-Weinberg equilibrium were excluded. Samples with a call rate < 95%, extreme heterozygosity, or of non-European ancestry based on genetic PCs were further excluded.

All genotype data were imputed to the 1000 Genomes Phase 3 version 5 reference panel separately by study and platform [26]. Number of individuals included in our GWAS by study and platform can be found in Additional file 1: Table S1.

### Association test

All four V assessments (V65L, V75L, V65H, and V75H) were standardized to have mean zero and unit standard deviation before analysis. SNP association analyses were performed within each study by platform for each of the four V assessments using linear regression assuming an additive dosage effect. RVtests [27] was used for NHS/NHSII cohorts (ALymetrix 6.0, Illumina HumanHap, Illumina OmniExpress, and Illumina OncoArray) and variants were removed from individual platform results if the expected minor allele counts were below 10. PLINK 2.0 [28] was used for MMHS cohorts (iCOGS and OncoArray). We ran six models adjusting for different covariates: Model 0 was the base model adjusting for age and the top four genetic PCs to account for population structure. Model 1 further adjusted for BMI. In addition to the covariates in Model 1, Model 2 further adjusted for PD, Model 3 further adjusted for DA, and Model 4 further adjusted for NDA. Model 5 was the fully adjusted model with age, BMI, genetic PCs, PD, DA, and NDA as covariates. Fixed effect meta-analyses across studies and platforms were conducted for each V assessment and model using METAL [29]. Cochran’s Q statistic was used to check for heterogeneity between studies and platforms. Quantile-quantile plots and genomic inflation factors were used to assess systematic inflation in test statistics due to population substructure. Manhattan plots were generated to visualize the overall GWAS results. LocusZoom plots [30] of the 1Mb region centered around the identified lead SNPs were generated to visualize the regional association results and nearby genes.

Given that the four V assessments were highly correlated with each other and might be proxies for an underlying latent phenotype, we performed multivariate phenotype association tests to pool association evidence across the four V assessments and get a single summary test statistic for each variant. We used R package MPAT [31] to obtain the summary *P* values and corresponding Z scores using test statistics from the meta-analysis results for each V assessment and model, accounting for sample overlaps of the four V assessments. We referred to this summary phenotype as VSUM, which was used as our primary multivariate outcome. SNPs with *P* < 5 × 10^−8^ in any of the six models for any of the four univariate V assessments or the multivariate VSUM were considered genome-wide significant.

### V, MD phenotypes, and breast cancer susceptibility variants

We evaluated whether the identified V loci were also associated with MD phenotypes or breast cancer risk using GWAS results from Breast Cancer Association Consortium (BCAC) [8, 25, 32]. To further identify shared susceptibility SNPs between V, MD phenotypes, and breast cancer risk, we conducted *in silico* lookups of 72 genome-wide significant MD phenotype SNPs identified by Sieh et al. [6] and Chen et al. [8], and 195 genome-wide significant breast cancer risk SNPs identified by Michailidou et al. [25] and Zhang et al. [32] in our GWAS of V. These candidate SNPs were considered significant for V if they passed the Bonferroni-corrected thresholds accounting for the number of MD (*P* < 0.05/72) or breast cancer (*P* < 0.05/195) SNPs tested in Model 0 for any V assessment.

### Genetic correlation and SNP-set test

Genetic correlations of the four V assessments and VSUM with MD phenotypes, breast cancer risk, overall and stratified by estrogen receptor (ER) status, adult BMI, childhood body fatness, age at menarche, and age at natural menopause were estimated using linkage disequilibrium (LD) score regression [33, 34]. Sources of summary statistics of these traits for estimating genetic correlations are summarized in Additional file 2: Table S2.

While genetic correlation quantifies the shared genetic contribution to two traits on genome-wide scale, it may also capture the contribution of other traits due to pleiotropy (e.g., the effect of BMI on the correlation between V and PD). Therefore, we further performed SNP-set tests to assess the genetic relationship between V and the above-mentioned traits using only reported genome-wide significant SNPs for those traits. SNPs for each trait were collected from published GWAS followed by LD clumping to remove any SNPs in LD (r^2^ > 0.1) with SNPs of smaller *P* value (see Additional file 2: Table S3). The test statistic for V and each trait was

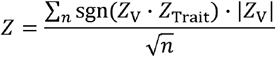

where z_V_ is the Z score from the SNP-specific association with V and z_Trait_ is the Z score from the SNP-specific association with the trait of interest, and n is the total number of tested genome-wide significant SNPs for that trait.

### Sensitivity analysis

Our study population contains both women who developed breast cancer and women who did not develop breast cancer during the follow-up period after mammogram collection. We therefore further adjusted for breast cancer case-control status in Model 5 to assess its impact on the genetic associations. We performed a multicollinearity check for the identified genome-wide significant SNPs for Model 5, where we adjusted for all three MD phenotypes, by calculating the variance inflation factor (VIF). To assess the potential impact of outliers on the association results at the identified GWAS loci, we calculated the studentized residuals for all samples for each genome-wide significant SNP. Samples with absolute studentized residual greater than 3 were considered as outliers.

## Results

Our GWAS meta-analysis of V comprised 7,040 women of European ancestry within the NHS, NHSII, and MMHS cohorts (Table 1). Women in MMHS were older, had higher BMI and lower MD compared to women in NHS and NHSII. Quantile-quantile plots and genomic inflation factors indicate there was no evidence of systematic inflation of the GWAS test statistics in any model for any V assessment (Additional file 2: Figure S1). Manhattan plots showing the -log_10_(*P*) for all tested SNPs across chromosomes are present in Additional file 2: Figure S2. Quantile-quantile plots of the heterogeneity *P* value indicate there was limited evidence of heterogeneity in the test results across studies and platforms (Additional file 2: Figure S3).

**Table 1.** Characteristics of NHS/NHSII and MMHS study population

|  | **NHS/NHSII (n = 4831)** | **MMHS (n = 2209)** |
| --- | --- | --- |
|  | Mean (SD) | Mean (SD) |
| **Age (years)** | 53.8 (9.2) | 58.9 (11.9) |
| **BMI (kg/m<sup>2</sup>)** | 25.9 (5.3) | 28.0 (6.2) |
| **Dense area** | 43.4 (29.3) | 23.8 (17.0) |
| **Nondense area** | 109.3 (73.7) | 130 (67.1) |
| **Percent density** | 32.8 (19.7) | 17.9 (12.9) |

*Abbreviations: NHS Nurses' Health Study, MMHS Mayo Mammography Health Study, SD standard deviation, BMI body mass index*

In total, we identified three independent loci that reached the genome-wide significant threshold of *P* < 5 × 10^−8^ in any model for any V assessment: 6q24.1 (*ECT2L*), 8q24.22 (*LINC01591*), and 12q22 (*PGAM1P5*) (Table 2). 6q24.1 (Lead SNP: rs138141444, *P* = 1.24 × 10^−8^ for V75H, Model 0) is a novel locus that has not previously been associated with MD phenotypes or breast cancer risk. Fig. 2a shows the regional association results for 6q24.1 from Model 0 (adjusting for age and genetic PCs) for V75H where the association was genome-wide significant. The association results were consistent across models with the same direction and similar effect sizes as well as *P* values. 8q24.22 (Lead SNP: rs79670367, *P* = 2.38 × 10^−8^ for VSUM, Model 5) is neither a MD nor breast cancer risk locus. Fig. 2b shows the regional association results for 8q24.22 from Model 5 (adjusting for age, BMI, DA, NDA, PD, and genetic PCs) for VSUM. The association between V and rs79670367 was more significant when we adjusted for PD (Model 2), DA (Model 3), or both (Model 5) and became less significant without adjustment for any MD phenotypes (Model 0 and 1) or adjusting for NDA only (Model 4). The direction of association was consistent across models. 12q22 (Lead SNP: rs113174754, *P* = 4.42 × 10^−8^ for VSUM, Model 3) has previously been associated with NDA (rs11836367, *P* = 8.40 × 10^−9^, r^2^ = 0.59 with rs113174754) [6], overall breast cancer risk (rs113174754, *P* = 1.08 × 10^−24^), and ER+ breast cancer risk (rs113174754, *P* = 1.37 × 10^−18^) [25]. This locus is also significantly associated with breast size (rs17356907, *P* = 1.30 × 10^−13^, r^2^ = 0.47 with rs113174754) [35]. Fig. 2c shows the regional association results for 12q22 from Model 3 (adjusting for age, BMI, DA, and genetic PCs) for VSUM. The association between V and rs113174754 became non-significant when we adjusted for NDA. The direction of association with V was consistent across models and consistent with the association with NDA (opposite direction) and breast cancer risk (same direction).

**Table 2.**
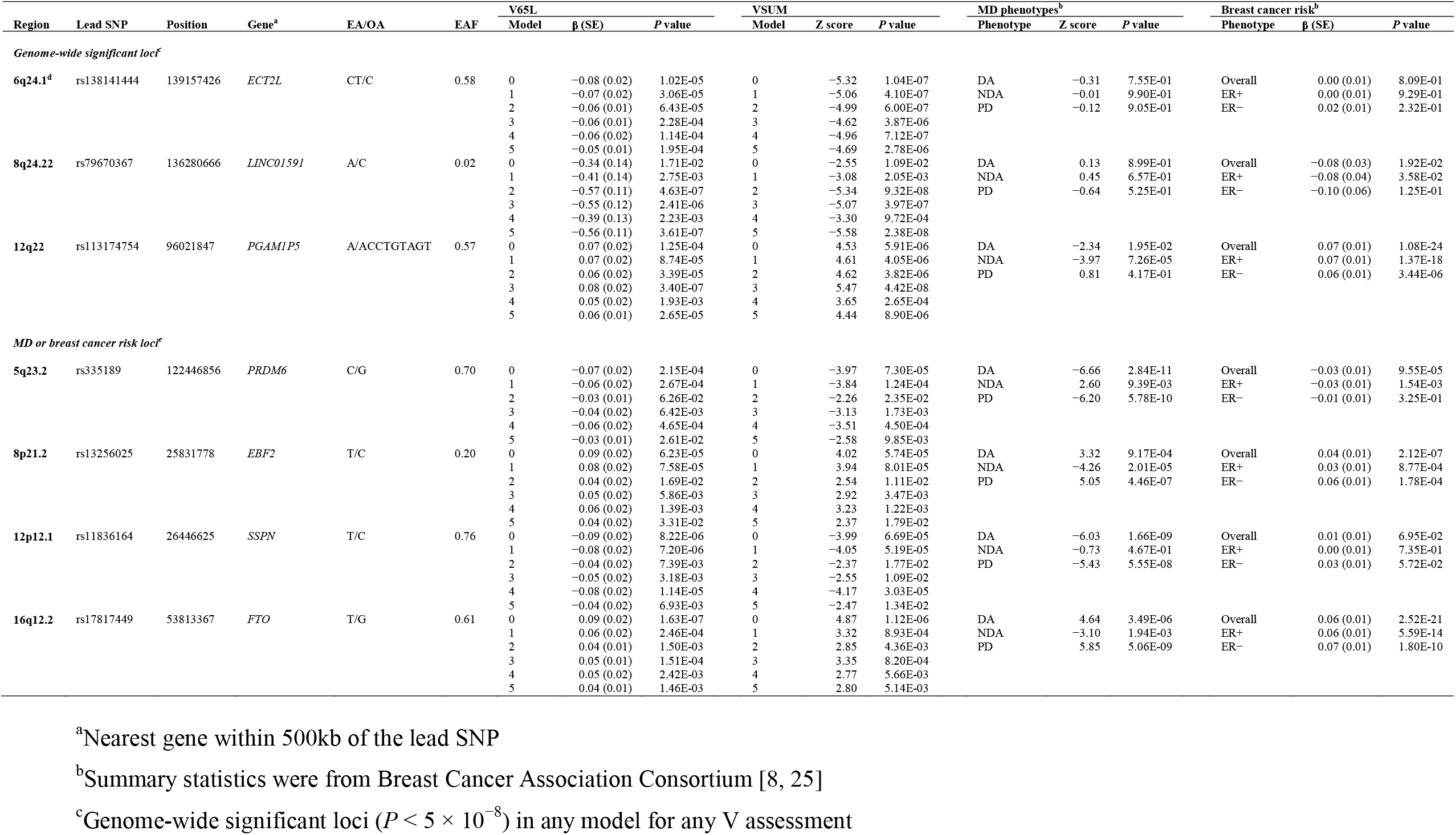
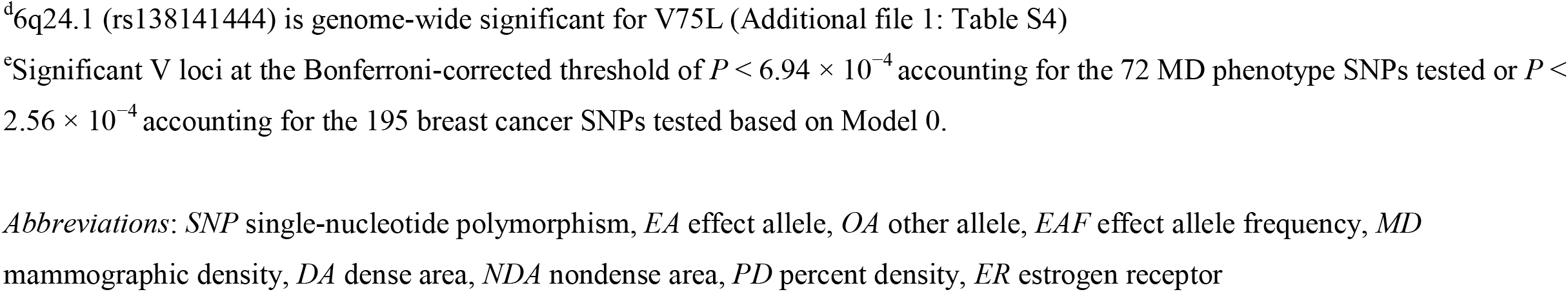
Novel V loci and their associations with mammographic density phenotypes and breast cancer risk, overall and stratified by estrogen receptor status

**Fig. 2.**
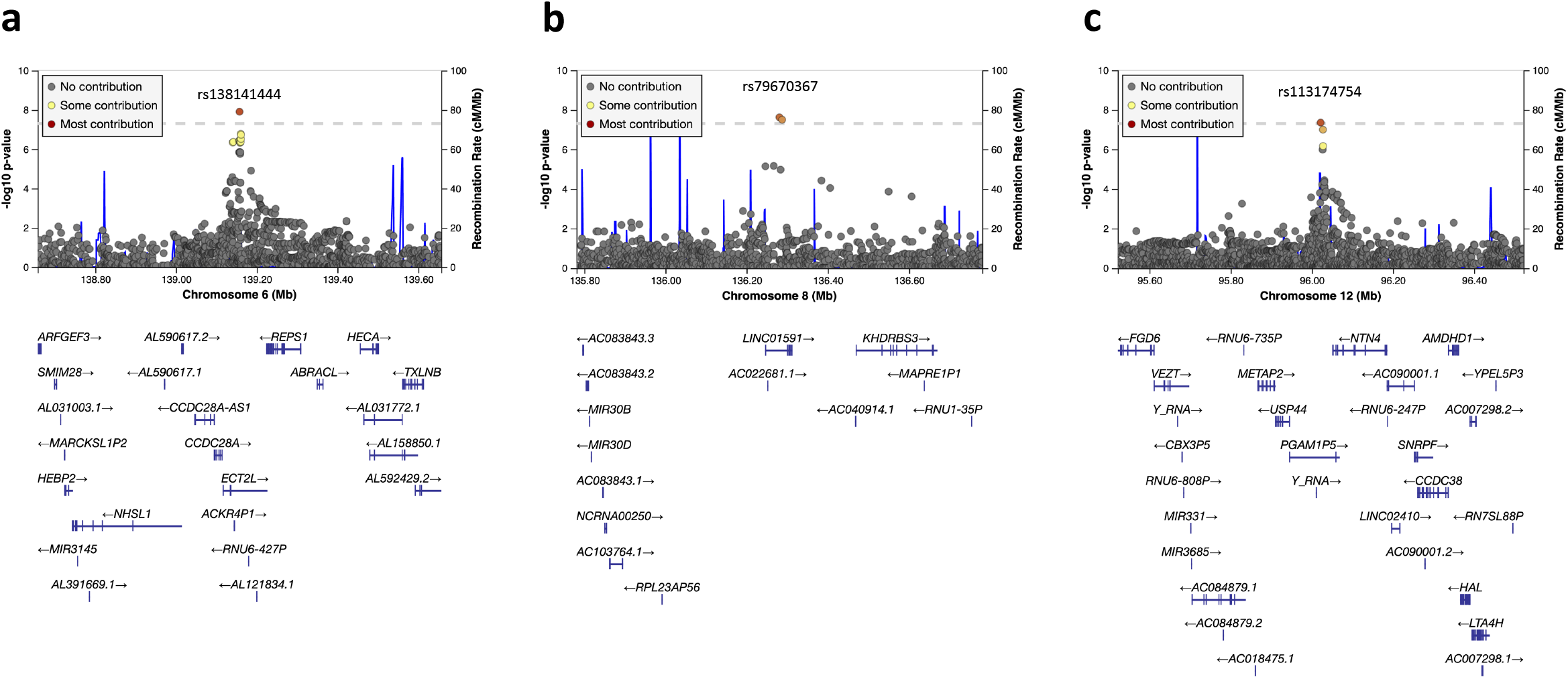
Regional association plots for the three genome-wide significant V loci. **a** = 6q24.1 (rs138141444; V75H, Model 0); **b** = 8q24.22 (rs79670367; VSUM, Model 5); **c** = 12q22 (rs113174754; VSUM, Model 3). Model and V assessment with the most significant results for each locus are shown. Each plot is centered around the lead SNP of each locus. SNPs in the 95% credible set at each locus are shown in color. Physical positions are based on NCBI Genome Reference Consortium Human Build 37. Plots were generated using LocusZoom [30]. *Abbreviations*: *SNP* single-nucleotide polymorphism

We identified four additional loci that had previously been associated with MD phenotypes or breast cancer risk and reached the Bonferroni-corrected thresholds accounting for the number of MD or breast cancer SNPs tested (*P* < 0.05/72 = 6.94 × 10^−4^ for MD, *P* < 0.05/195 = 2.56 × 10^−4^ for breast cancer risk) in Model 0: 5q23.2 (*PRDM6*), 8p21.2 (*EBF2*), 12p12.1 (*SSPN*), and 16q12.2 (*FTO*) (Table 2). 5q23.2 (Lead SNP: rs335189, *P* = 7.30 × 10^−5^ for VSUM, Model 0) is a known locus for DA (*P* = 2.84 × 10^−11^) and PD (*P* = 5.78 × 10^−10^) [8]. The associations with V were significant in Model 0, Model 1 (adjusting for age, BMI, and genetic PCs), and Model 4 (adjusting for age, BMI, NDA, and genetic PCs) but not with adjustment for DA, PD, or both (Model 2, 3, and 5). 8p21.2 (Lead SNP: rs13256025, *P* = 5.74 × 10^−5^ for VSUM, Model 0) has previously been associated with breast cancer risk (*P* = 1.40 × 10^−8^) [32]. The associations with V were significant in Model 0 and 1, and became non-significant when we adjusted for DA, NDA, or PD (Model 2, 3, 4, and 5). Although this locus has not been reported as a MD locus, the *P* value of the association between the lead SNP and PD was close to the genome-wide significant threshold (*P* = 4.46 × 10^−7^) [8]. 12p12.1 (Lead SNP: rs11836164, *P* = 6.69 × 10^−5^ for VSUM, Model 0) is a known locus for DA (*P* = 1.66 × 10^−9^) [8]. The associations with V were significant in Model 0, 1, and 4, and became non-significant when we adjusted for DA, PD, or both (Model 2, 3, and 5). 16q12.2 (Lead SNP: rs17817449, *P* = 1.12 × 10^−6^ for VSUM, Model 0) is a known locus for PD (*P* = 5.06 × 10^−9^) [8], overall (*P* = 2.52 × 10^−21^), ER+ (*P* = 5.59 × 10^−14^), and ER-breast cancer risk (*P* = 1.80 × 10^−10^). This locus is also significantly associated with BMI (rs17817449, *P* = 5.10 × 10^−19^) [36] and breast size (rs62033406, *P* = 3.70 × 10^−7^, r^2^ = 0.89 with rs17817449) [35]. The associations with V were significant in Model 0 and became non-significant when we adjusted for BMI or any MD phenotype. The directions of association with V were consistent with those significant associations with MD (same direction for PD and DA, opposite direction for NDA) or breast cancer (same direction) for all four loci. Association results of all identified V loci for all models and V assessments can be found in Additional file 1: Table S4. There was no substantial difference between the results of different V assessments. The full lookup results of the 72 MD phenotype SNPs and 195 breast cancer SNPs can be found in Additional file 1: Table S5 and Table S6.

We observed significant positive genetic correlations between V and dense area (r_g_ = 0.79, *P* = 5.91 × 10^−5^ for VSUM, Model 0) and percent density (r_g_ = 0.73, *P* = 1 × 10^−4^ for VSUM, Model 0) (Fig. 3a). The correlations became non-significant using GWAS results from Model 2 (adjusting for age, BMI, PD, and genetic PCs). Positive correlations were also observed with overall (r_g_ = 0.20, *P* = 6.90 × 10^−3^ for VSUM, Model 0) and ER+ (r_g_ = 0.22, *P* = 4.60 × 10^−3^ for VSUM, Model 0) breast cancer and became non-significant when adjusting for PD. We also observed a significant negative association with adult BMI (r_g_ = -0.36, *P* = 3.88 × 10^−7^ for VSUM, Model 0), which became non-significant when adjusting for BMI. A strong negative correlation was observed for NDA (r_g_ = -0.60, *P* = 5.20 × 10^−3^ for VSUM, Model 0) before adjusting for PD. Genetic correlation results were similar across V assessments; the full results are summarized in Additional file 1: Table S7.

**Fig. 3.**
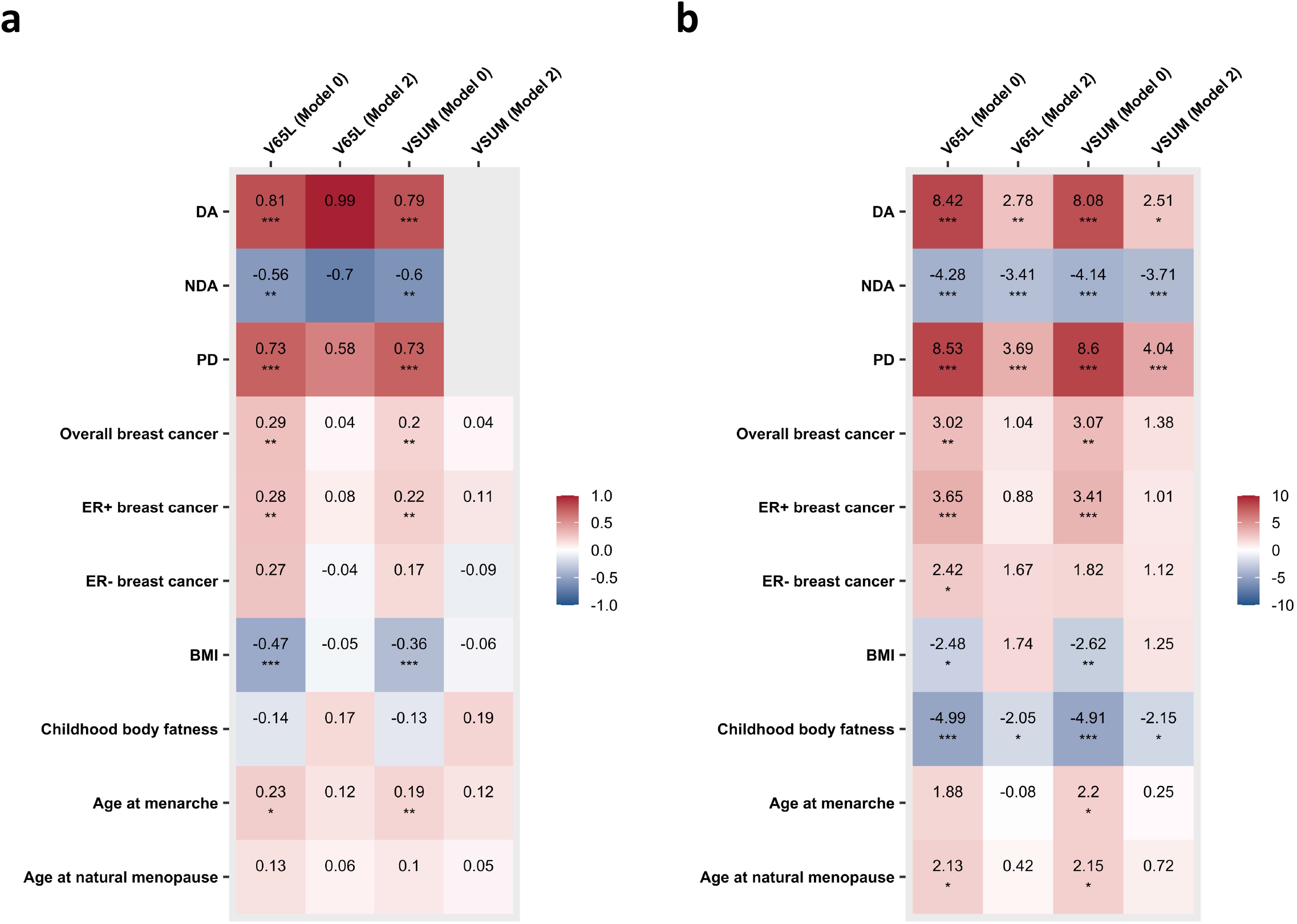
Genetic correlation and SNP-set test results of V with mammographic density phenotypes, breast cancer risk, and other breast cancer risk factors. **a** = genetic correlations between V and other traits; **b** = SNP-set test results of the relationship of V and other traits. Results of Model 0 and 2 for V65L and VSUM are shown. Estimates passed the Bonferroni threshold (*P* < 0.05/40 = 1.25 × 10^−3^) are marked with triple asterisk (***); estimates with *P* < 0.01 are marked with double asterisk (**); estimates with nominal significance (*P* < 0.05) are marked with single asterisk (*). Genetic correlations between VSUM (Model 2) and MD phenotypes were not estimated due to the out of bounds heritability of V. *Abbreviations*: *SNP* single-nucleotide polymorphism, *DA* dense area, *NDA* nondense area, *PD* percent density, *ER* estrogen receptor, *BMI* body mass index

In addition to the genetic relationships of V with DA, NDA, PD, and breast cancer risk identified by genetic correlations, we further identified a significant positive association between V and ER+ breast cancer (z = 3.41, *P* = 6.41 × 10^−4^ for VSUM, Model 0) and a significant negative association between V and childhood body fatness from the SNP-set test using genome-wide significant SNPs for childhood body fatness (z = -4.91, *P* = 9.05 × 10^−7^ for VSUM, Model 0) (Fig. 3b). The overall pattern of the associations was similar for genetic correlation and SNP-set test. It is worth noting that for MD phenotypes and childhood body fatness, the associations with V remained nominally significant (*P* < 0.05) if we further adjust for BMI and PD in the SNP-set test. Plots showing Z scores from GWAS of V and GWAS of MD phenotypes [8] and breast cancer [25] for the SNPs included in the SNP-set tests are present in Fig. 4. SNP-set test results across all models and V assessments can be found in Additional file 1: Table S8.

**Fig. 4.**
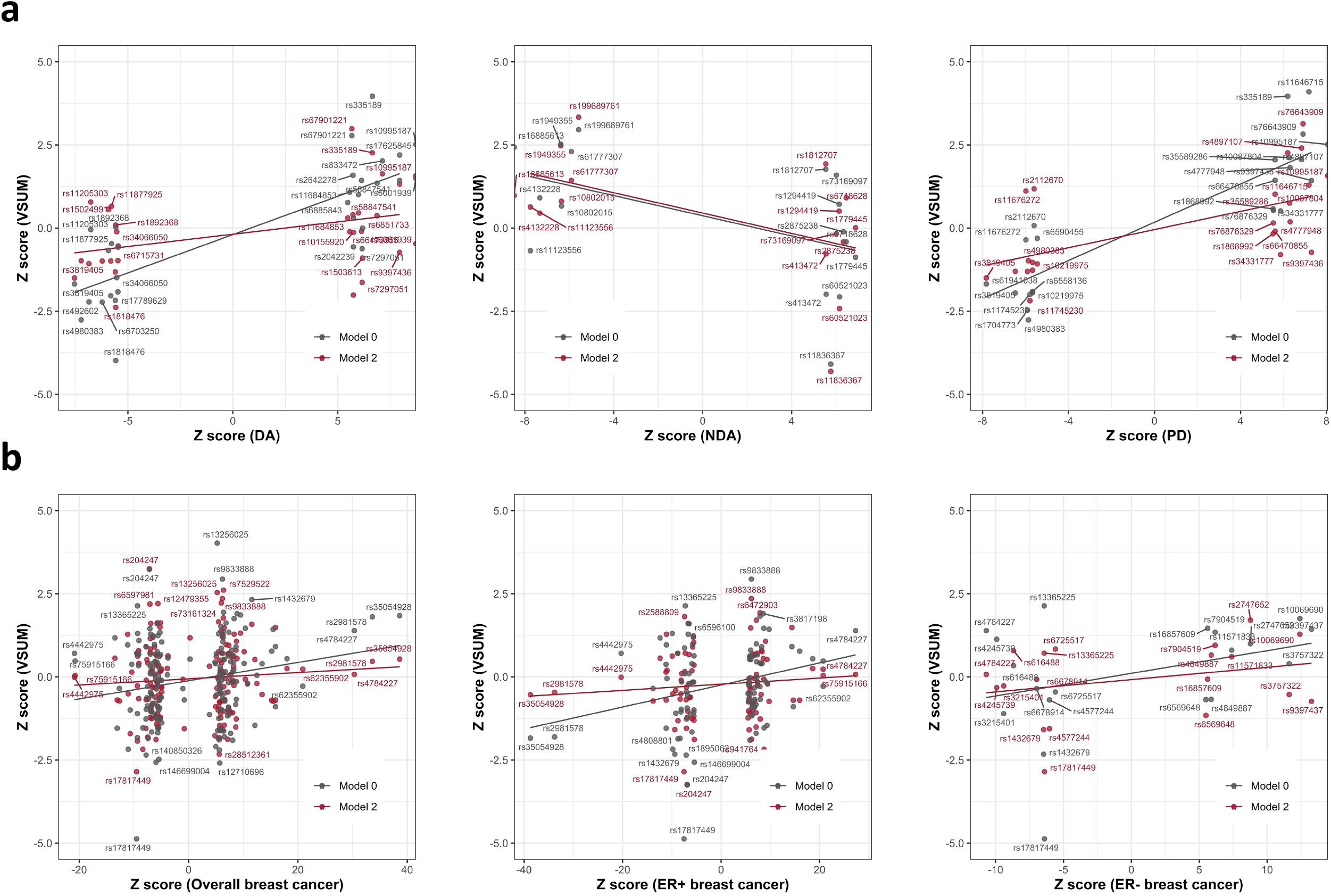
Z scores from GWAS of V, mammographic density phenotypes, and breast cancer risk for SNPs included in SNP-set test. **a** = scatter plots of Z scores from GWAS of V by Z scores from GWAS of percent density (PD), dense area (DA), and nondense area (NDA) for mammographic density SNPs; **b** = scatter plots of Z scores from GWAS of V by Z scores from GWAS of overall breast cancer risk and stratified by estrogen receptor (ER) status for breast cancer SNPs. For each SNP, GWAS results from Model 0 and 2 for VSUM are shown with gray and red dots, respectively. RS number for some SNPs are not shown on the plots. Gray line is the fitted linear regression line of Z scores for results from Model 0; red line is the fitted linear regression line of Z scores for results from Model 2. Note that some of the overall breast cancer risk SNPs are not genome-wide significant because we obtained the Z scores from one study and those SNPs were reported by other studies. *Abbreviations*: *GWAS* genome-wide association study, *SNP* single-nucleotide polymorphism

No substantial change on the top findings was observed after including breast cancer case-control status as a covariate (Additional file 1: Table S4). There was no multicollinearity issue for the effect estimates of the genome-wide significant SNPs in Model 5 (VIFs all close to 1). There were 10 outliers with absolute studentized residual greater than 3 for rs79670367 at 8q24.22 from Model 5 for V65L. The effect estimates for the effect allele increased by 24% after removing those outliers. No substantial impact of outliers was found for other identified V SNPs.

## Discussion

While MD continues to be one of the most well-established and widely used mammographic risk factors for breast cancer, there are gaps in our knowledge of mammographic features themselves and their relationship with breast cancer risk. Current MD measures do not capture the heterogeneity in the distribution of dense breast tissue on a mammogram, known as texture variation. Increasing evidence have shown that the performance of texture variation on discriminating breast cancer outcomes is either comparable or even higher than the performance of MD measures [12, 16, 37, 38]. Understanding the contributing mechanisms of texture variation on breast cancer risk, especially the involved genetic components, would expand our knowledge on breast cancer development. In this study, we performed the first GWAS meta-analysis of mammographic texture variation, focusing on a summary measure of gray scale variation on mammograms (V). We identified three genome-wide significant V loci: 6q24.1 (*ECT2L*), 8q24.22 (*LINC01591*), and 12q22 (*PGAM1P5*), the first two of which have not previously been associated with MD or breast cancer risk. Four additional loci for MD or breast cancer risk, 5q23.2 (*PRDM6*), 8p21.2 (*EBF2*), 12p12.1 (*SSPN*), and 16q12.2 (*FTO*), were also found associated with V.

Different models of the SNP-V association were fit to capture different effects. Model 0 with only age and genetic PCs as covariates can capture both the effect of genetic variants on V and the effect that was mediated by BMI or MD phenotypes. We also fit Model 5 adjusting for all MD phenotypes together to assess the variant effect that was independent of all adjusted covariates. Although PD can be calculated from DA and NDA, previous GWAS of MD still identified different loci and genetic effects for different MD measures. We therefore fit the fully adjusted Model 5 to minimize the effect of MD phenotypes on the V associations. Collinearity issue in Model 5 did not have an impact on the effect estimates of the variants. Comparing the results from different models may also provide evidence for the underlying relationships between the genetic variants, V, and other adjusted covariates as well as boost power to detect V SNPs.

For example, if we observed a SNP-V association in models with and without adjustment for MD, then it is likely that the SNP influences V through other pathways that are independent of density; if the SNP-V association was only observed in model without adjusting for density, then it indicates that the SNP effect on V might be largely mediated by density. Downstream analyses need to be performed to confirm the relationships. Both V65L and the calculated summary statistics of the four V assessments, VSUM, were used as our primary outcomes. We have a larger sample size thus a greater power for low resolution V assessments compared to high resolution assessments (sample size for V65L and V75L = 7,040; sample size for V65H and V75H = 4,763). Although a previous study looking at the relationship between V and breast cancer risk in NHS/NHSII used a different assessment, V75L, as the outcome [16], these two low resolution V assessments were highly correlated with each other (ρ = 0.98, Fig. 1) and there was no substantial difference in the GWAS results of these two assessments (Additional file 1: Table S4). Using VSUM also has the advantage of boosting power given that the SNP associations were similar across different V assessments.

Among the three genome-wide significant V loci, **12q22** is also associated with NDA and breast cancer risk in consistent direction, suggesting that at least part of its genetic effect on V is mediated by NDA or the genetic effect on NDA is mediated by V, and there are potential shared biological pathways between these three traits. These hypotheses are further supported by the fact that 12q22 is also associated with total breast size and its association with V was most significant when adjusting for DA and became non-significant when adjusting for NDA. The lead variant rs113174754 at 12q22 is an indel near pseudogene *PGAM1P5* and is 30kb upstream of protein coding gene *NTN4* (see Fig. 2c). *NTN4* encodes a member of the netrin family of proteins, which involved in axon guidance, tumorigenesis, and angiogenesis. NDA SNP at 12q22 (rs11836367-C, correlated with the effect allele of rs113174754) has been found to downregulate *NTN4* in mammary tissue [6]. *NTN4* has also been identified as a candidate breast cancer risk gene by colocalization analysis, where the C allele of SNP rs61938093 (r^2^ = 0.48 with the effect allele of rs113174754) at this region reduced *NTN4* promoter activity and knockdown of *NTN4* promoted breast cell proliferation and tumor growth [39]. These findings suggest a shared genetic basis and potential biological mechanisms for mammographic risk factors, especially breast adipose tissue (represented by NDA), and breast cancer risk at this locus, and may also explain the observed association between V and breast cancer risk. **6q24.1** and **8q24.22** are V loci that have not been seen associated with MD phenotypes or breast cancer risk. The lead variant rs138141444 at 6q24.1 is an intronic indel in *ECT2L*. The lead variant rs79670367 at 8q24.22 is an intronic SNP in *LINC01591*. Neither these two genes nor nearby genes have been associated with breast cancer risk. The genetic effects of these two loci on V are therefore likely through mechanisms not mediated by MD. It should also be noted that the effect allele frequency for rs79670367 is less than 5% and the outlier analysis indicated that the association results might be influenced by influential outliers. Moreover, only about half of the samples have genotype data on this variant (available in NHS/NHSII Illumina HumanHap and MMHS OncoArray). Further studies are needed to confirm the findings at these two loci.

Four additional V loci have previously been associated with breast cancer risk or MD phenotypes. The lead variant rs13256025 at **8p21.2** is an intronic SNP in protein coding gene *EBF2. EBF2* encodes well conserved DNA-binding helix-loop-helix transcription factors, which involved in differentiation of osteoblasts. Although little is known about the role of *EBF2* in breast cancer development, studies have shown that inactivation of *EBF* genes can lead to tumorigenesis via accumulation and expansion of undifferentiated progenitor cells [40]. **16q12.2** is associated with both PD and breast cancer risk in the same direction with its lead SNP rs17817449 located in *FTO. FTO* is a well-established susceptibility gene for obesity [41]. In our analysis, the association was only significant in the base model and became non-significant when adjusting for BMI, suggesting that its genetic effect on V might be mediated by BMI. *FTO* is overexpressed in breast cancer cells, which affects the energy metabolism of the cells [42]. **5q23.2** is a known locus for DA and PD. The lead variant rs335189 is an intronic SNP in *PRDM6. PRDM6* encodes a transcriptional repressor involved in the regulation of endothelial cell proliferation, survival, and differentiation, and may play a role in breast cancer tumorigenesis [7, 43]. The lead variant rs11836164 at **12p12.1** is an intronic SNP near *SSPN* and is only associated with DA. Functional analysis needs to be performed to further investigate the role of identified V SNPs in mammary development and breast cancer etiology.

Consistent with the phenotypic relationships we observed for V and MD measures, there were strong positive genetic correlations of V with DA and PD, and negative genetic correlations with NDA. The positive genetic correlations between V and breast cancer risk (overall and ER+ specific) were also nominally significant, further supporting that the observed phenotypic association between V and breast cancer risk can at least be partially explained by shared genetic components. The magnitude of these genetic correlations is comparable to those between MD and breast cancer risk [6]. A genetic variant can be associated with multiple traits, which is known as pleiotropy. Studies have shown that jointly analyzing GWAS data of multiple traits can boost power to detect genetic associations for each trait and improve the prediction performance [44, 45]. In our analysis, we observed significant genetic correlations of V with MD phenotypes and BMI using genome-wide association results. It is therefore very likely that a substantial number of variants are associated with both MD phenotypes, especially NDA, and BMI, which would dilute the correlations we observed for any pair of the traits. SNP-set tests may provide more evidence for the shared mechanism underlying two traits using only susceptibility variants. Here, we found that even if we adjust for PD in the model, there were still significant correlations between V and PD based on genome-wide significant SNPs for PD, indicating that the genetic contribution of V cannot be fully explained by PD and PD is either a mediator or collider of the association between the genetic variants and V (Fig. 4a). Correlations of V with breast cancer and childhood body fatness were also stronger at the susceptibility variants. There were still correlations, though not significant, after adjusting for PD, providing evidence for the genetic relationship between V and these traits that were not mediated by MD (Fig. 4b).

Our study focuses on a summary texture measure, V, but there are also many other texture features. For example, Manduca et al. systematically evaluated 1,443 textural features and identified six independently validated strongest features [13]. Malkov et al. identified 15 texture features that were significantly associated with breast cancer risk, several of which were only weakly correlated with PD [46]. Studying the genetics of these features or their combinations may provide additional information for the genetic architecture of breast parenchymal texture variation. Our study included breast cancer cases, which might be concerning since V has been associated with breast cancer risk. However, both theoretical [47] and empirical [48] evidence suggest that including cases of a rare outcome does not bias the association estimates in GWAS of a secondary outcome, except when both the genetic variant being analyzed and the secondary outcome are very strong risk factors—stronger than those exhibited by breast cancer risk SNPs, V, or BMI. Indeed, we did not observe any substantial changes on the top findings after further adjusting for breast cancer case-control status in the model. Moreover, the direction of the associations we observed—e.g., a breast cancer risk allele was positively associated with V—are opposite of those expected if the SNP-V association is solely an artefact due to collider bias.

Multiple testing issue caused by studying four V assessments may also be a concern, we therefore estimated a single summary test statistic, VSUM, to minimize the impact of multiple testing and to boost power. Studying the computerized automated texture feature can also reduce the potential bias caused by measurement error that studies on semi-automated MD measures are usually susceptible to.

## Conclusions

In conclusion, we performed a GWAS of breast parenchymal texture variation, V, and identified three independent loci at genome-wide significance, including 12q22 (*PGAM1P5*) that are associated with MD phenotypes and breast cancer risk, and 6q24.1 (*ECT2L*) and 8q24.22 (*LINC01591*) that are novel V susceptibility loci. Four additional V loci were identified from looking up MD and breast cancer susceptibility SNPs in GWAS of V, including 5q23.2 (*PRDM6*), 8p21.2 (*EBF2*), 12p12.1 (*SSPN*), and 16q12.2 (*FTO*). These findings provide the first evidence of the genetic basis of V and shared genetic components between V, MD, and breast cancer risk. Future studies are needed to confirm our findings and further improve our understanding of the mechanisms underlying the relationship between texture features, MD, and breast cancer development.

## Data Availability

NHS/NHSII: The data that support the findings of this study are available from the Nurses' Health Studies, however they are not publicly available. Investigators interested in using the data can request access, and feasibility will be discussed at an investigators' meeting. Limits are not placed on scientific questions or methods, and there is no requirement for co-authorship. Additional data sharing information and policy details can be accessed at http://www.nurseshealthstudy.org/researchers.
MMHS: The summary statistics generated from the current study are available from the corresponding author on reasonable request.

## List of abbreviations

BCAC: Breast Cancer Association Consortium
BMI: body mass index
DA: dense area
ER: estrogen receptor
GWAS: genome-wide association study
LD: linkage disequilibrium
MD: mammographic density
MMHS: Mayo Mammography Health Study
NDA: nondense area
NHS: Nurses’ Health Study
PC: principal component
PD: percent density
SNP: single-nucleotide polymorphism
VIF: variance inflation factor
V75H: V with 25% erosion and high resolution
V75L: V with 25% erosion and low resolution
V65H: V with 35% erosion and high resolution
V65L: V with 35% erosion and low resolution

## Declarations

### Ethics approval and consent to participate

The study protocol was approved by the institutional review boards of the Brigham and Women’s Hospital, Harvard T.H. Chan School of Public Health, and the Mayo Clinic, and was in accordance with the 1964 Declaration of Helsinki and its later amendments or comparable ethical standards. All participants provided written informed consent.

### Consent for publication

Not applicable

### Availability of data and materials

NHS/NHSII: The data that support the findings of this study are available from the Nurses’ Health Studies, however they are not publicly available. Investigators interested in using the data can request access, and feasibility will be discussed at an investigators’ meeting. Limits are not placed on scientific questions or methods, and there is no requirement for co-authorship. Additional data sharing information and policy details can be accessed at http://www.nurseshealthstudy.org/researchers.

MMHS: The summary statistics generated from the current study are available from the corresponding author on reasonable request.

### Authors’ contributions

XJ, PK, RMT, and CVM conceived and designed the study. RMT and ETW prepared the mammographic texture variation data for NHS/NHSII. CMV and JH prepared the mammographic texture variation data for MMHS. CT prepared the genotype data for NHS/NHSII. SJW prepared the genotype data for MMHS. HC and SL prepared the mammographic density GWAS data. YL analyzed and interpreted the data, and was a major contributor in writing the manuscript. All authors read and approved the final manuscript.

## Acknowledgements

We would like to thank the participants and staff of the NHS and NHSII for their valuable contributions as well as the following state cancer registries for their help: AL, AZ, AR, CA, CO, CT, DE, FL, GA, ID, IL, IN, IA, KY, LA, ME, MD, MA, MI, NE, NH, NJ, NY, NC, ND, OH, OK, OR, PA, RI, SC, TN, TX, VA, WA, WY.

The BCAC MD GWAS was supported by CA244670 and CA194393. BCAC is funded by the European Union’s Horizon 2020 Research and Innovation Programme (grant numbers 634935 and 633784 for BRIDGES and B-CAST respectively), and the PERSPECTIVE I&I project, funded by the Government of Canada through Genome Canada and the Canadian Institutes of Health Research, the Ministère de l’Économie et de l’Innovation du Québec through Genome Québec, the Quebec Breast Cancer Foundation. The EU Horizon 2020 Research and Innovation Programme funding source had no role in study design, data collection, data analysis, data interpretation or writing of the report. Additional funding for BCAC is provided via the Confluence project which is funded with intramural funds from the National Cancer Institute Intramural Research Program, National Institutes of Health.

Genotyping of the OncoArray was funded by the NIH Grant U19 CA148065, and Cancer UK Grant C1287/A16563 and the PERSPECTIVE project supported by the Government of Canada through Genome Canada and the Canadian Institutes of Health Research (grant GPH-129344) and, the Ministère de l’Économie, Science et Innovation du Québec through Genome Québec and the PSRSIIRI-701 grant, and the Quebec Breast Cancer Foundation. Funding for iCOGS came from: the European Community’s Seventh Framework Programme under grant agreement n° 223175 (HEALTH-F2-2009-223175) (COGS), Cancer Research UK (C1287/A10118, C1287/A10710, C12292/A11174, C1281/A12014, C5047/A8384, C5047/A15007, C5047/A10692, C8197/A16565), the National Institutes of Health (CA128978) and Post-Cancer GWAS initiative (1U19 CA148537, 1U19 CA148065 and 1U19 CA148112 - the GAME-ON initiative), the Department of Defence (W81XWH-10-1-0341), the Canadian Institutes of Health Research (CIHR) for the CIHR Team in Familial Risks of Breast Cancer, and Komen Foundation for the Cure, the Breast Cancer Research Foundation, and the Ovarian Cancer Research Fund.

## Additional files

### Additional file 1 (XLSX)

**Table S1**. Number of GWAS subjects by study and platform.

**Table S4**. GWAS results of significant V loci for all V assessments and models.

**Table S5**. Lookup results of 72 MD phenotype SNPs in GWAS of V.

**Table S6**. Lookup results of 195 breast cancer SNPs in GWAS of V.

**Table S7**. Genetic correlation results of all V assessments and models.

**Table S8**. SNP-set test results of all V assessments and models.

### Additional file 2 (DOCX)

**Table S2**. Sources of summary statistics of breast cancer risk and breast cancer risk factors for calculating genetic correlation.

**Table S3**. Sources of summary statistics of breast cancer risk and breast cancer risk factors for the SNP-set test.

**Figure S1**. Quantile-quantile plots of the GWAS meta-analysis results.

**Figure S2**. Manhattan plots of the GWAS meta-analysis results.

**Figure S3**. Quantile-quantile plots of the *P* value of heterogeneity.

## References

1. Byrne C, Schairer C, Wolfe J, Parekh N, Salane M, Brinton LA, Hoover R, Haile R: Mammographic features and breast cancer risk: effects with time, age, and menopause status. J Natl Cancer Inst 1995, 87(21):1622–1629.

2. Boyd NF, Martin LJ, Yaffe MJ, Minkin S: Mammographic density and breast cancer risk: current understanding and future prospects. Breast Cancer Res 2011, 13(6):223.

3. Boyd NF, Guo H, Martin LJ, Sun L, Stone J, Fishell E, Jong RA, Hislop G, Chiarelli A, Minkin S et al: Mammographic density and the risk and detection of breast cancer. N Engl J Med 2007, 356(3):227–236.

4. Boyd NF, Dite GS, Stone J, Gunasekara A, English DR, McCredie MR, Giles GG, Tritchler D, Chiarelli A, Yaffe MJ et al: Heritability of mammographic density, a risk factor for breast cancer. N Engl J Med 2002, 347(12):886–894.

5. Stone J, Dite GS, Gunasekara A, English DR, McCredie MR, Giles GG, Cawson JN, Hegele RA, Chiarelli AM, Yaffe MJ et al: The heritability of mammographically dense and nondense breast tissue. Cancer Epidemiol Biomarkers Prev 2006, 15(4):612–617.

6. Sieh W, Rothstein JH, Klein RJ, Alexeeff SE, Sakoda LC, Jorgenson E, McBride RB, Graff RE, McGuire V, Achacoso N et al: Identification of 31 loci for mammographic density phenotypes and their associations with breast cancer risk. Nat Commun 2020, 11(1):5116.

7. Lindstrom S, Thompson DJ, Paterson AD, Li J, Gierach GL, Scott C, Stone J, Douglas JA, dos-Santos-Silva I, Fernandez-Navarro P et al: Genome-wide association study identifies multiple loci associated with both mammographic density and breast cancer risk. Nat Commun 2014, 5:5303.

8. Chen H, Fan S, Stone J, Thompson DJ, Douglas J, Li S, Scott C, Bolla MK, Wang Q, Dennis J et al: Genome-wide and transcriptome-wide association studies of mammographic density phenotypes reveal novel loci. Breast Cancer Res 2022, 24(1):27.

9. Boyd NF, Rommens JM, Vogt K, Lee V, Hopper JL, Yaffe MJ, Paterson AD: Mammographic breast density as an intermediate phenotype for breast cancer. Lancet Oncol 2005, 6(10):798–808.

10. Byng JW, Boyd NF, Fishell E, Jong RA, Yaffe MJ: The quantitative analysis of mammographic densities. Phys Med Biol 1994, 39(10):1629–1638.

11. Gastounioti A, Conant EF, Kontos D: Beyond breast density: a review on the advancing role of parenchymal texture analysis in breast cancer risk assessment. Breast Cancer Res 2016, 18(1):91.

12. Heine JJ, Scott CG, Sellers TA, Brandt KR, Serie DJ, Wu FF, Morton MJ, Schueler BA, Couch FJ, Olson JE et al: A novel automated mammographic density measure and breast cancer risk. J Natl Cancer Inst 2012, 104(13):1028–1037.

13. Manduca A, Carston MJ, Heine JJ, Scott CG, Pankratz VS, Brandt KR, Sellers TA, Vachon CM, Cerhan JR: Texture features from mammographic images and risk of breast cancer. Cancer Epidemiol Biomarkers Prev 2009, 18(3):837–845.

14. Nielsen M, Karemore G, Loog M, Raundahl J, Karssemeijer N, Otten JD, Karsdal MA, Vachon CM, Christiansen C: A novel and automatic mammographic texture resemblance marker is an independent risk factor for breast cancer. Cancer Epidemiol 2011, 35(4):381–387.

15. Wanders JOP, van Gils CH, Karssemeijer N, Holland K, Kallenberg M, Peeters PHM, Nielsen M, Lillholm M: The combined effect of mammographic texture and density on breast cancer risk: a cohort study. Breast Cancer Res 2018, 20(1):36.

16. Warner ET, Rice MS, Zeleznik OA, Fowler EE, Murthy D, Vachon CM, Bertrand KA, Rosner BA, Heine J, Tamimi RM: Automated percent mammographic density, mammographic texture variation, and risk of breast cancer: a nested case-control study. NPJ Breast Cancer 2021, 7(1):68.

17. Oh H, Rice MS, Warner ET, Bertrand KA, Fowler EE, Eliassen AH, Rosner BA, Heine JJ, Tamimi RM: Early-Life and Adult Anthropometrics in Relation to Mammographic Image Intensity Variation in the Nurses’ Health Studies. Cancer Epidemiol Biomarkers Prev 2020, 29(2):343–351.

18. Tworoger SS, Missmer SA, Eliassen AH, Spiegelman D, Folkerd E, Dowsett M, Barbieri RL, Hankinson SE: The association of plasma DHEA and DHEA sulfate with breast cancer risk in predominantly premenopausal women. Cancer Epidemiol Biomarkers Prev 2006, 15(5):967–971.

19. Olson JE, Sellers TA, Scott CG, Schueler BA, Brandt KR, Serie DJ, Jensen MR, Wu FF, Morton MJ, Heine JJ et al: The influence of mammogram acquisition on the mammographic density and breast cancer association in the Mayo Mammography Health Study cohort. Breast Cancer Res 2012, 14(6):R147.

20. Heine JJ, Cao K, Rollison DE: Calibrated measures for breast density estimation. Acad Radiol 2011, 18(5):547–555.

21. Heine JJ, Cao K, Rollison DE, Tiffenberg G, Thomas JA: A quantitative description of the percentage of breast density measurement using full-field digital mammography. Acad Radiol 2011, 18(5):556–564.

22. Boyd NF, Stone J, Martin LJ, Jong R, Fishell E, Yaffe M, Hammond G, Minkin S: The association of breast mitogens with mammographic densities. Br J Cancer 2002, 87(8):876–882.

23. Yaghjyan L, Pettersson A, Colditz GA, Collins LC, Schnitt SJ, Beck AH, Rosner B, Vachon C, Tamimi RM: Postmenopausal mammographic breast density and subsequent breast cancer risk according to selected tissue markers. Br J Cancer 2015, 113(7):1104–1113.

24. Lindstrom S, Loomis S, Turman C, Huang H, Huang J, Aschard H, Chan AT, Choi H, Cornelis M, Curhan G et al: A comprehensive survey of genetic variation in 20,691 subjects from four large cohorts. PLoS One 2017, 12(3):e0173997.

25. Michailidou K, Lindstrom S, Dennis J, Beesley J, Hui S, Kar S, Lemacon A, Soucy P, Glubb D, Rostamianfar A et al: Association analysis identifies 65 new breast cancer risk loci. Nature 2017, 551(7678):92–94.

26. Sudmant PH, Rausch T, Gardner EJ, Handsaker RE, Abyzov A, Huddleston J, Zhang Y, Ye K, Jun G, Fritz MH et al: An integrated map of structural variation in 2,504 human genomes. Nature 2015, 526(7571):75–81.

27. Zhan X, Hu Y, Li B, Abecasis GR, Liu DJ: RVTESTS: an efficient and comprehensive tool for rare variant association analysis using sequence data. Bioinformatics 2016, 32(9):1423–1426.

28. Chang CC, Chow CC, Tellier LC, Vattikuti S, Purcell SM, Lee JJ: Second-generation PLINK: rising to the challenge of larger and richer datasets. Gigascience 2015, 4:7.

29. Willer CJ, Li Y, Abecasis GR: METAL: fast and efficient meta-analysis of genomewide association scans. Bioinformatics 2010, 26(17):2190–2191.

30. Boughton AP, Welch RP, Flickinger M, VandeHaar P, Taliun D, Abecasis GR, Boehnke M: LocusZoom.js: Interactive and embeddable visualization of genetic association study results. Bioinformatics 2021.

31. Liu Z, Lin X: A Geometric Perspective on the Power of Principal Component Association Tests in Multiple Phenotype Studies. J Am Stat Assoc 2019, 114(527):975–990.

32. Zhang H, Ahearn TU, Lecarpentier J, Barnes D, Beesley J, Qi G, Jiang X, O’Mara TA, Zhao N, Bolla MK et al: Genome-wide association study identifies 32 novel breast cancer susceptibility loci from overall and subtype-specific analyses. Nat Genet 2020, 52(6):572–581.

33. Bulik-Sullivan B, Finucane HK, Anttila V, Gusev A, Day FR, Loh PR, ReproGen C, Psychiatric Genomics C, Genetic Consortium for Anorexia Nervosa of the Wellcome Trust Case Control C, Duncan L et al: An atlas of genetic correlations across human diseases and traits. Nat Genet 2015, 47(11):1236–1241.

34. Bulik-Sullivan BK, Loh PR, Finucane HK, Ripke S, Yang J, Schizophrenia Working Group of the Psychiatric Genomics C, Patterson N, Daly MJ, Price AL, Neale BM: LD Score regression distinguishes confounding from polygenicity in genome-wide association studies. Nat Genet 2015, 47(3):291–295.

35. Pickrell JK, Berisa T, Liu JZ, Segurel L, Tung JY, Hinds DA: Detection and interpretation of shared genetic influences on 42 human traits. Nat Genet 2016, 48(7):709–717.

36. Wojcik GL, Graff M, Nishimura KK, Tao R, Haessler J, Gignoux CR, Highland HM, Patel YM, Sorokin EP, Avery CL et al: Genetic analyses of diverse populations improves discovery for complex traits. Nature 2019, 570(7762):514–518.

37. Gierach GL, Li H, Loud JT, Greene MH, Chow CK, Lan L, Prindiville SA, Eng-Wong J, Soballe PW, Giambartolomei C et al: Relationships between computer-extracted mammographic texture pattern features and BRCA1/2 mutation status: a cross-sectional study. Breast Cancer Res 2014, 16(4):424.

38. Li H, Giger ML, Olopade OI, Margolis A, Lan L, Chinander MR: Computerized texture analysis of mammographic parenchymal patterns of digitized mammograms. Acad Radiol 2005, 12(7):863–873.

39. Beesley J, Sivakumaran H, Moradi Marjaneh M, Shi W, Hillman KM, Kaufmann S, Hussein N, Kar S, Lima LG, Ham S et al: eQTL Colocalization Analyses Identify NTN4 as a Candidate Breast Cancer Risk Gene. Am J Hum Genet 2020, 107(4):778–787.

40. Liao D: Emerging roles of the EBF family of transcription factors in tumor suppression. Mol Cancer Res 2009, 7(12):1893–1901.

41. Loos RJ, Yeo GS: The bigger picture of FTO: the first GWAS-identified obesity gene. Nat Rev Endocrinol 2014, 10(1):51–61.

42. Liu Y, Wang R, Zhang L, Li J, Lou K, Shi B: The lipid metabolism gene FTO influences breast cancer cell energy metabolism via the PI3K/AKT signaling pathway. Oncol Lett 2017, 13(6):4685–4690.

43. Casamassimi A, Rienzo M, Di Zazzo E, Sorrentino A, Fiore D, Proto MC, Moncharmont B, Gazzerro P, Bifulco M, Abbondanza C: Multifaceted Role of PRDM Proteins in Human Cancer. Int J Mol Sci 2020, 21(7).

44. Aguirre M, Tanigawa Y, Venkataraman GR, Tibshirani R, Hastie T, Rivas MA: Polygenic risk modeling with latent trait-related genetic components. Eur J Hum Genet 2021, 29(7):1071–1081.

45. Turley P, Walters RK, Maghzian O, Okbay A, Lee JJ, Fontana MA, Nguyen-Viet TA, Wedow R, Zacher M, Furlotte NA et al: Multi-trait analysis of genome-wide association summary statistics using MTAG. Nat Genet 2018, 50(2):229–237.

46. Malkov S, Shepherd JA, Scott CG, Tamimi RM, Ma L, Bertrand KA, Couch F, Jensen MR, Mahmoudzadeh AP, Fan B et al: Mammographic texture and risk of breast cancer by tumor type and estrogen receptor status. Breast Cancer Res 2016, 18(1):122.

47. Monsees GM, Tamimi RM, Kraft P: Genome-wide association scans for secondary traits using case-control samples. Genet Epidemiol 2009, 33(8):717–728.

48. Lindstrom S, Vachon CM, Li J, Varghese J, Thompson D, Warren R, Brown J, Leyland J, Audley T, Wareham NJ et al: Common variants in ZNF365 are associated with both mammographic density and breast cancer risk. Nat Genet 2011, 43(3):185–187.

