## Additional File 2 for "A genome-wide association study of mammographic texture variation"

**Table S2.** Sources of summary statistics of breast cancer risk and breast cancer risk factors for calculating genetic correlation

| **Trait** | **Source** | **URL** |
| --- | --- | --- |
| Dense area | Chen et al. [1] | <https://bcac.ccge.medschl.cam.ac.uk/bcacdata/oncoarray/oncoarray-and-combined-summary-result/gwas-summary-results-mammographic-density-2021/> |
| Nondense area | Chen et al. [1] | <https://bcac.ccge.medschl.cam.ac.uk/bcacdata/oncoarray/oncoarray-and-combined-summary-result/gwas-summary-results-mammographic-density-2021/> |
| Percent density | Chen et al. [1] | <https://bcac.ccge.medschl.cam.ac.uk/bcacdata/oncoarray/oncoarray-and-combined-summary-result/gwas-summary-results-mammographic-density-2021/> |
| Overall breast cancer | Michailidou et al. [2] | <http://bcac.ccge.medschl.cam.ac.uk/bcacdata/oncoarray/oncoarray-and-combined-summary-result/gwas-summary-results-breast-cancer-risk-2017/> |
| ER+ breast cancer | Michailidou et al. [2] | <http://bcac.ccge.medschl.cam.ac.uk/bcacdata/oncoarray/oncoarray-and-combined-summary-result/gwas-summary-results-breast-cancer-risk-2017/> |
| ER− breast cancer | Michailidou et al. [2] | <http://bcac.ccge.medschl.cam.ac.uk/bcacdata/oncoarray/oncoarray-and-combined-summary-result/gwas-summary-results-breast-cancer-risk-2017/> |
| Adult body mass index | Pulit et al. [3] | <https://zenodo.org/record/1251813#.YPCd1xNKiSg> |
| Childhood body fatness | Warner et al. [4] | Not publicly available |
| Age at menarche | Day et al. [5] | <https://www.reprogen.org/data_download.html> |
| Age at natural menopause | Day et al. [6] | <https://www.reprogen.org/data_download.html> |

**Table S3.** Sources of summary statistics of breast cancer risk and breast cancer risk factors for the SNP-set test

| **Trait** | **Source** | **Number of SNPs included** |
| --- | --- | --- |
| Dense area | Sieh et al. [7] and Chen et al. [1] | 31 |
| Nondense area | Sieh et al. [7] and Chen et al. [1] | 16 |
| Percent density | Sieh et al. [7] and Chen et al. [1] | 23 |
| Overall breast cancer | Michailidou et al. [2] and Zhang et al. [8] | 178 |
| ER+ breast cancer | Michailidou et al. [2] and Zhang et al. [8] | 101 |
| ER− breast cancer | Michailidou et al. [2] and Zhang et al. [8] | 27 |
| Adult body mass index | Pulit et al. [3] | 279 |
| Childhood body fatness | Warner et al. [4] | 18 |
| Age at menarche | Day et al. [5] | 377 |
| Age at natural menopause | Day et al. [6] | 54 |

**Figure S1.** Quantile-quantile plots of the GWAS meta-analysis results

**a**

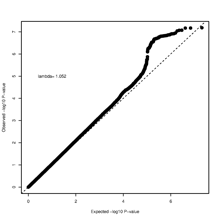

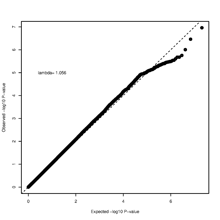

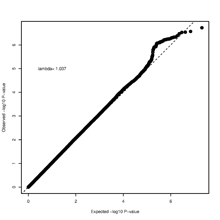

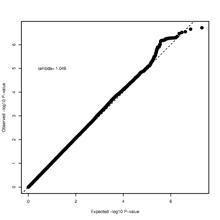

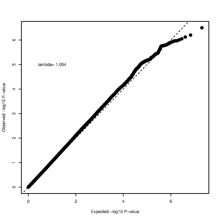

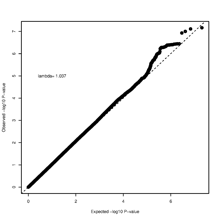

**b**

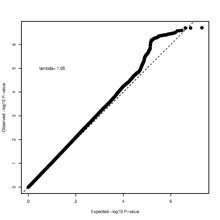

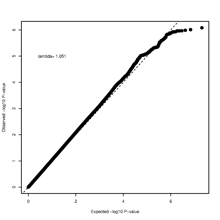

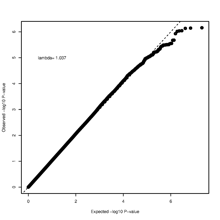

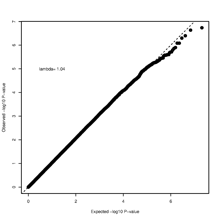

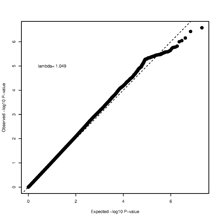

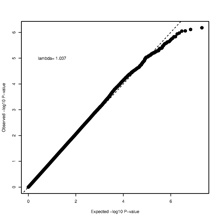

**c**

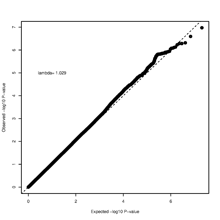

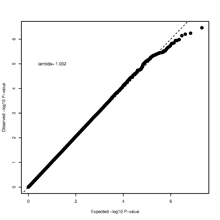

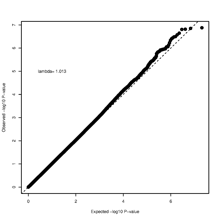

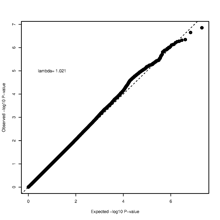

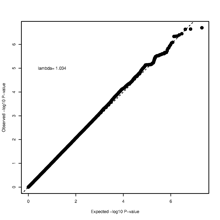

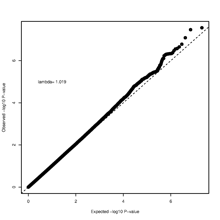

**d**

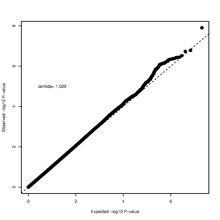

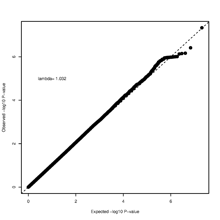

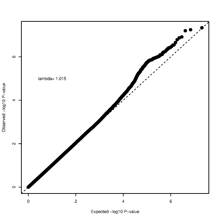

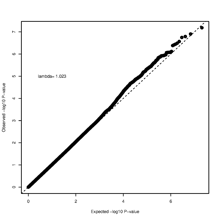

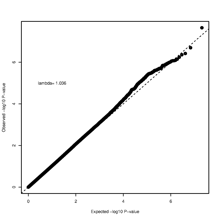

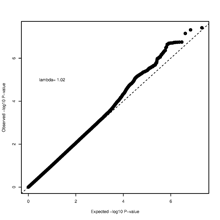

**e**

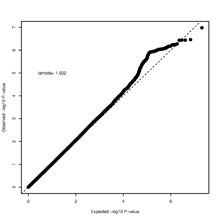

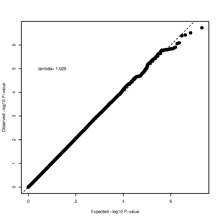

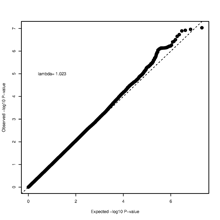

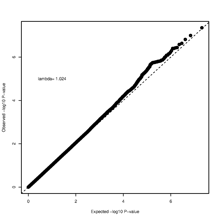

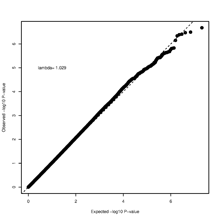

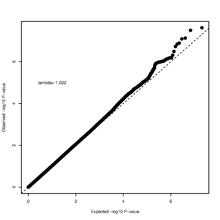

Quantile-quantile plots of the GWAS meta-analysis results. **a** = V65L; **b** = V75L; **c** = V65H; **d** = V75H; **e** = VSUM. On each panel, the plots corresponding to Model 0 to Model 5 from left to right.

**Figure S2.** Manhattan plots of the GWAS meta-analysis results

**a**

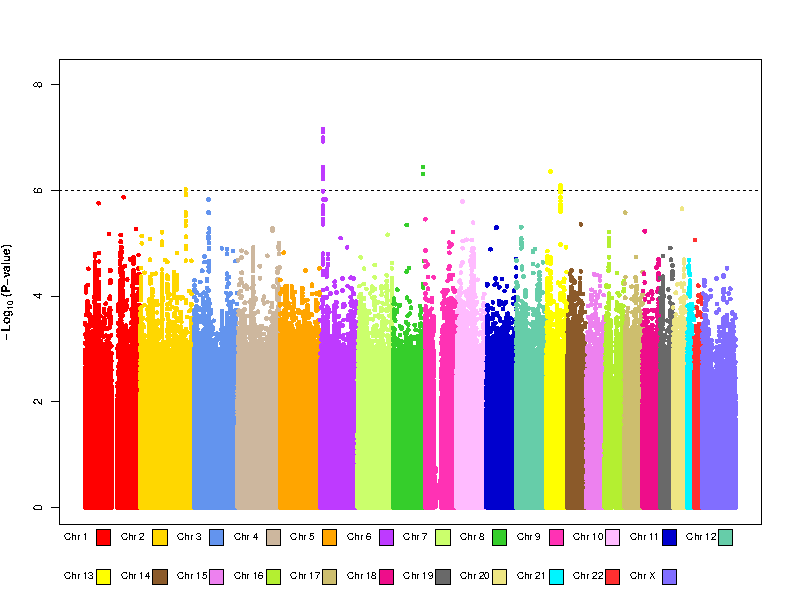

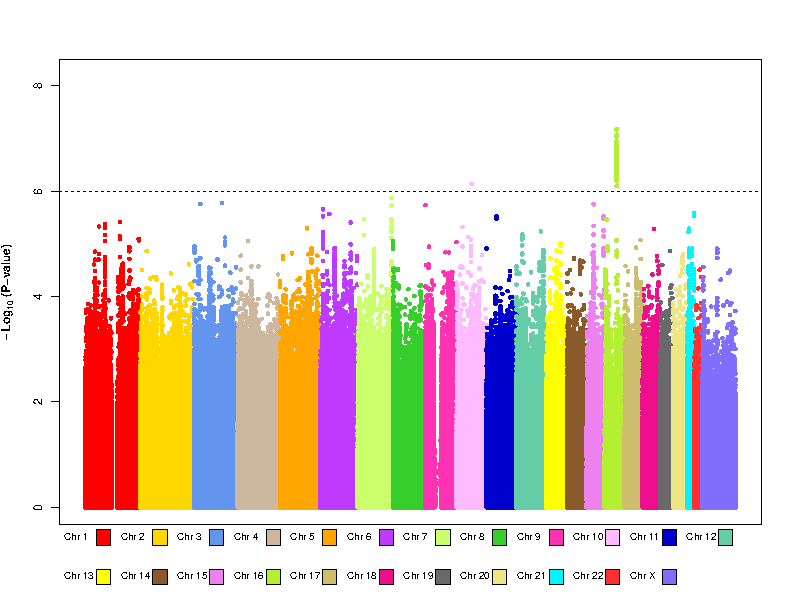

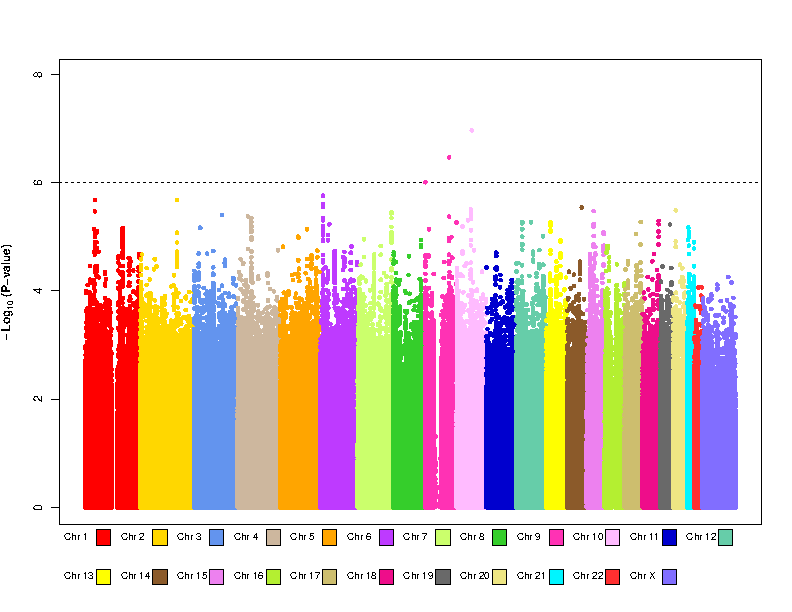

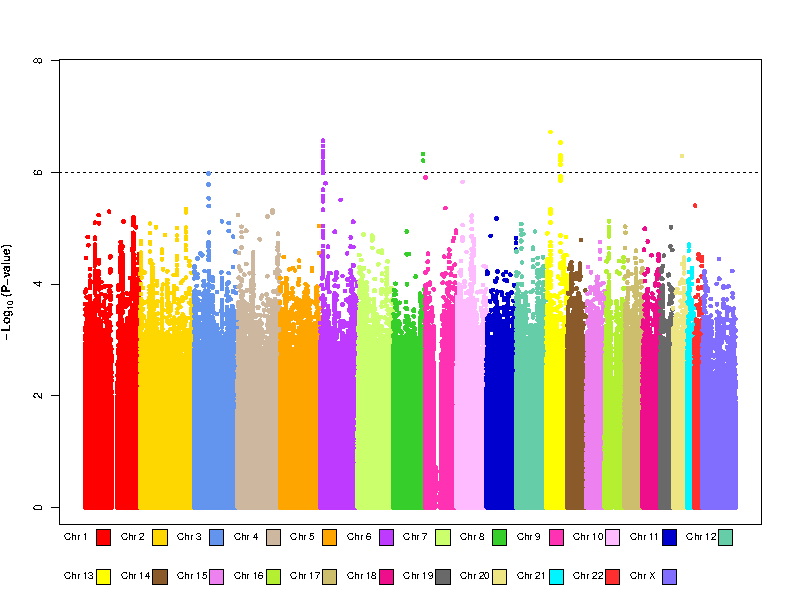

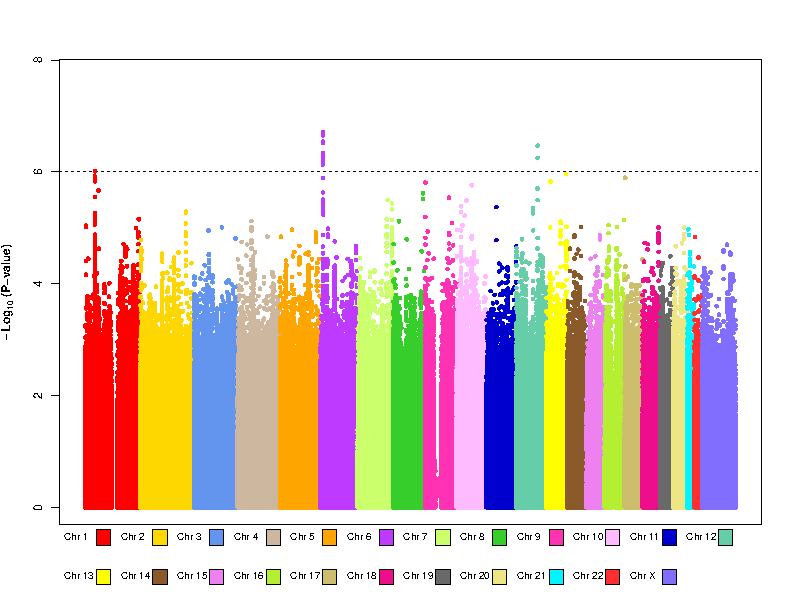

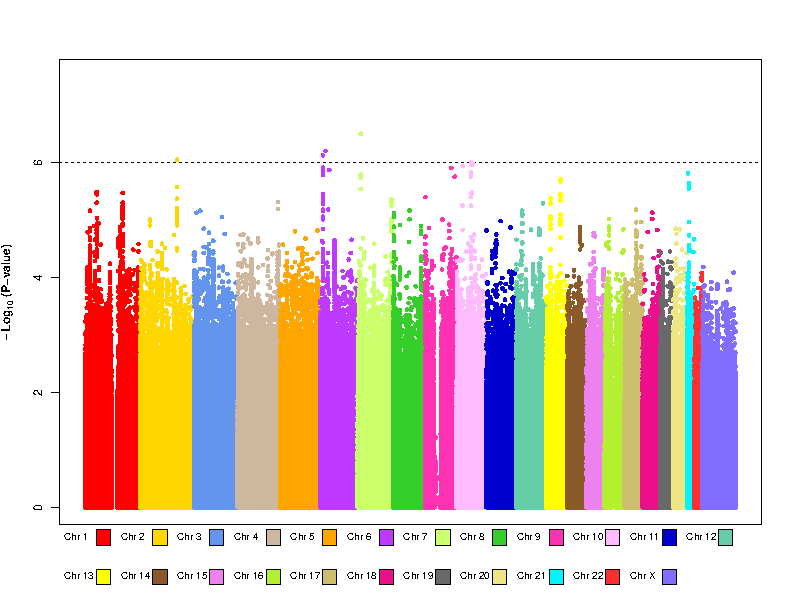

**b**

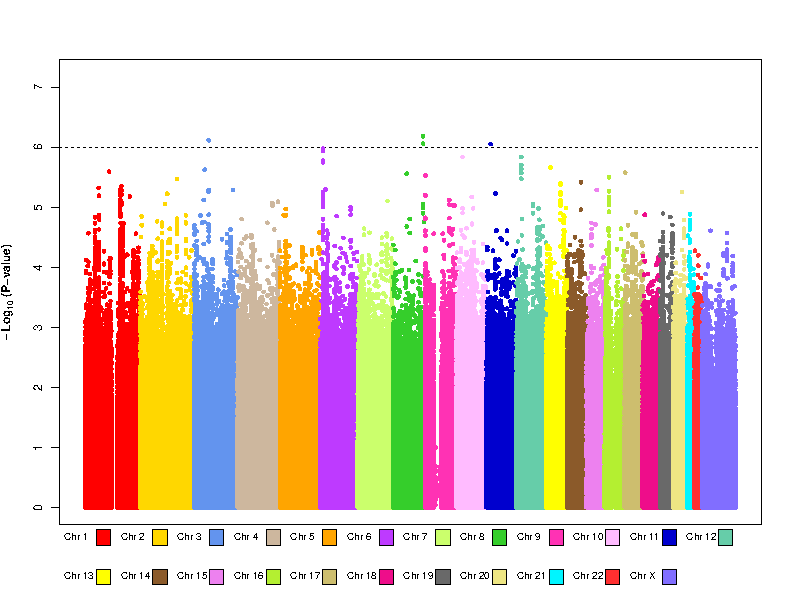

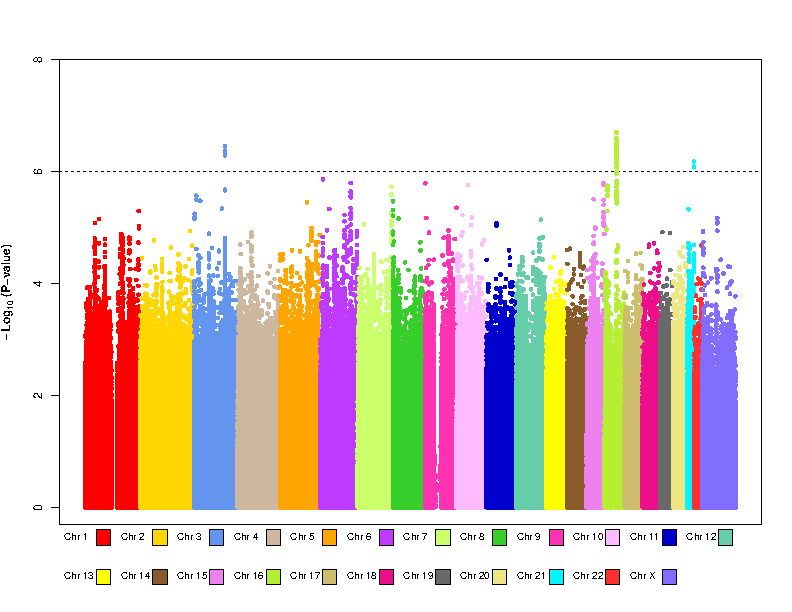

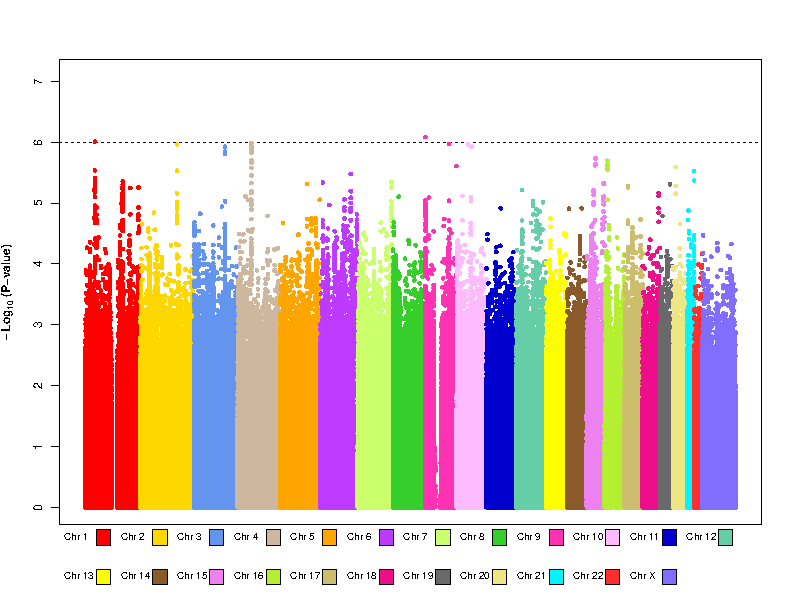

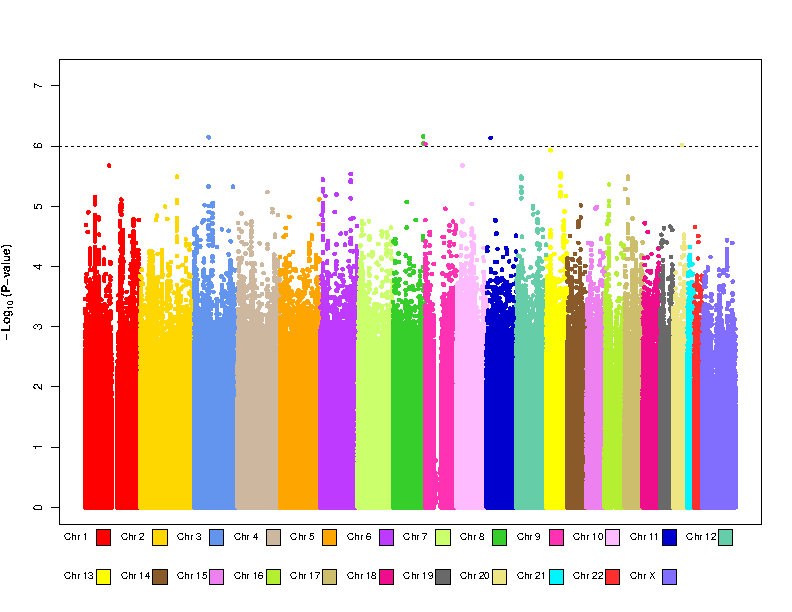

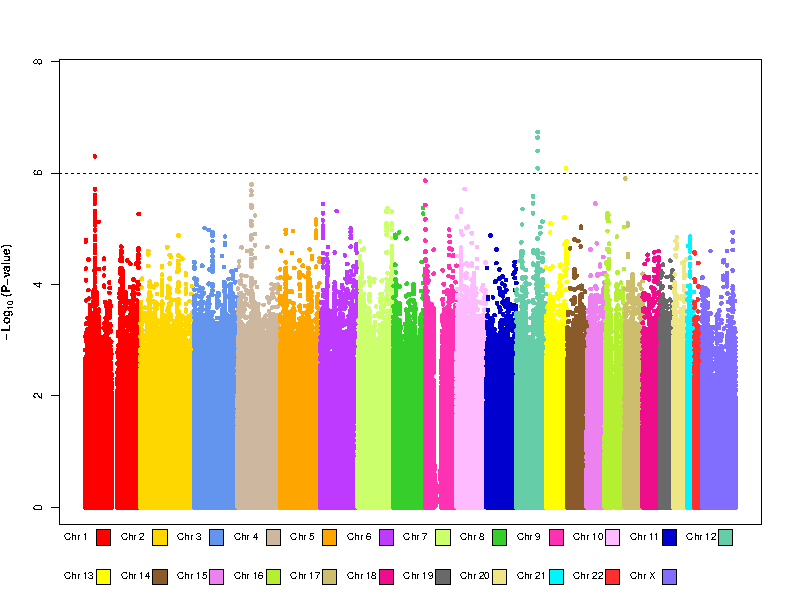

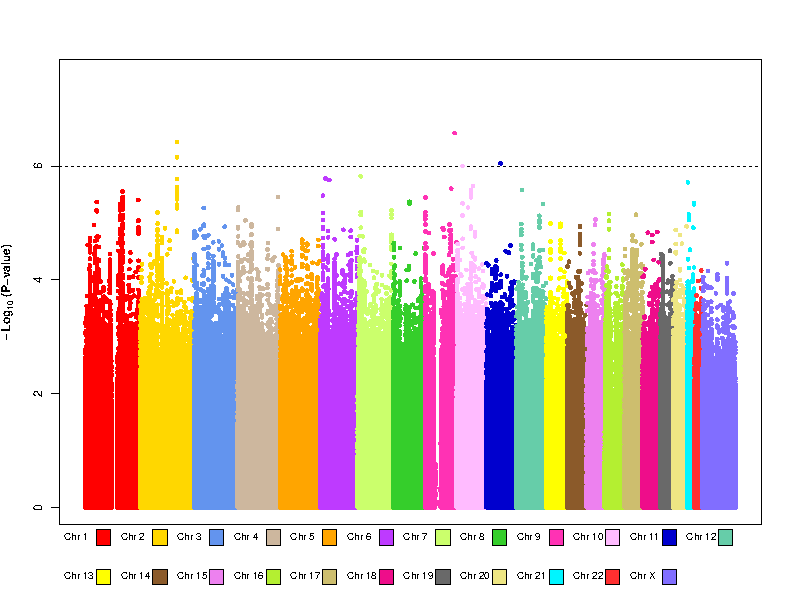

**c**

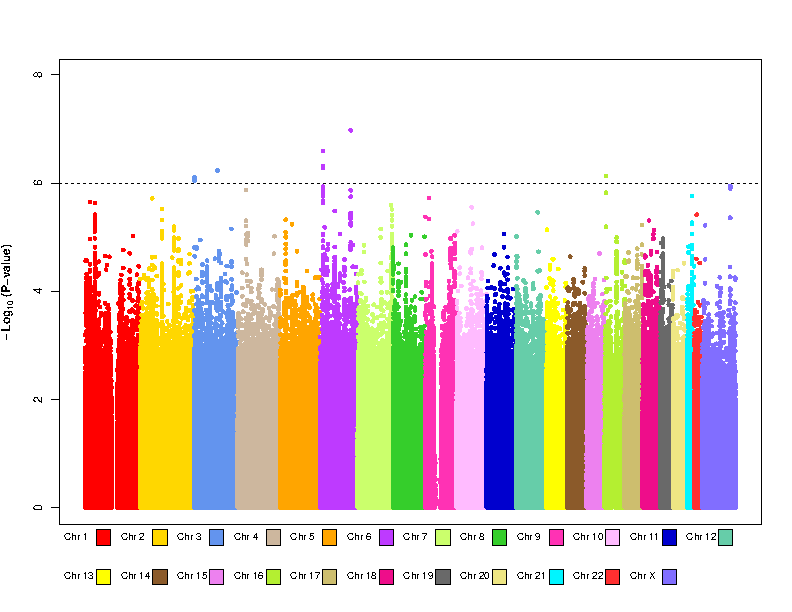

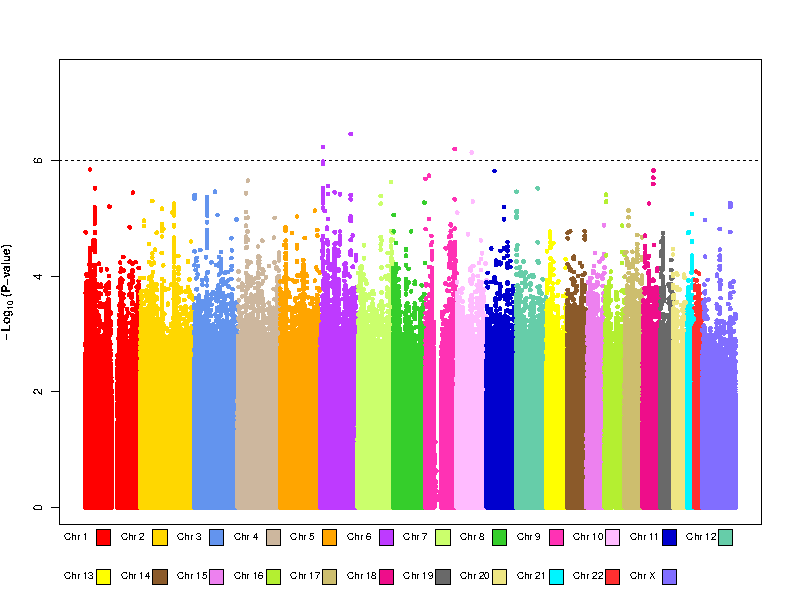

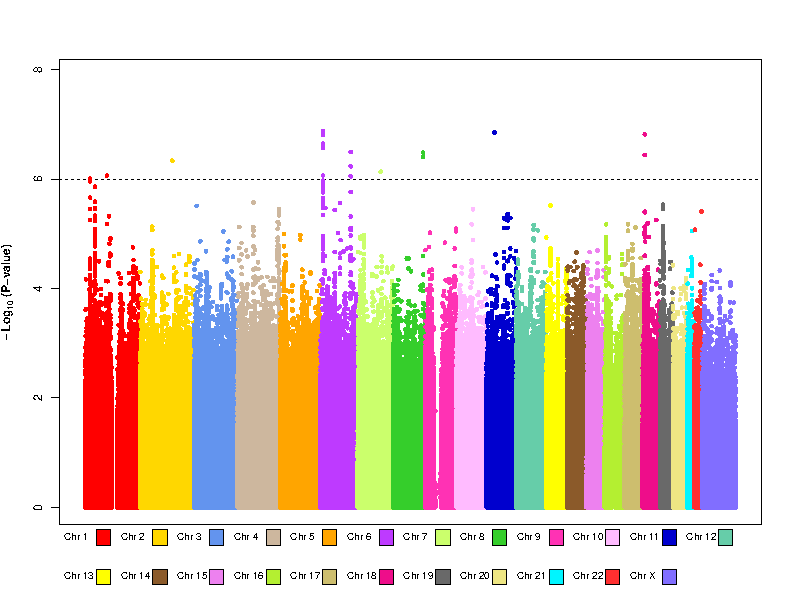

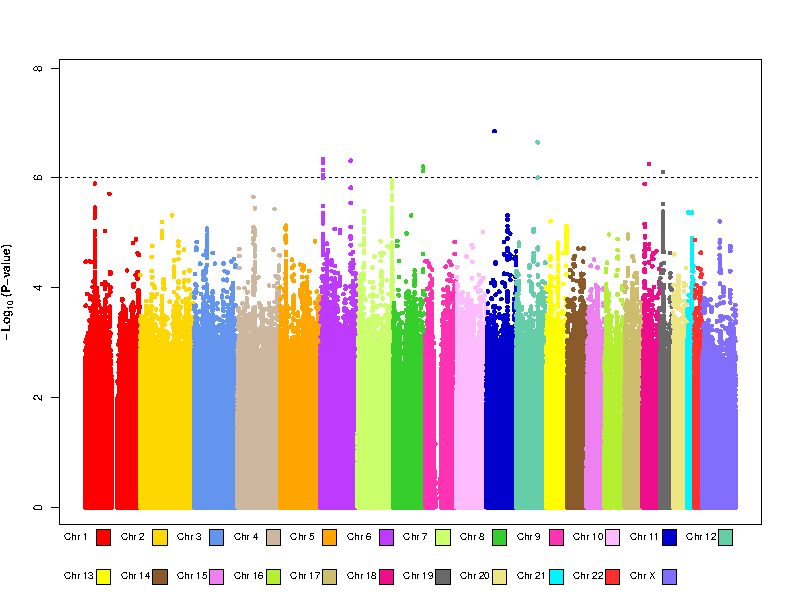

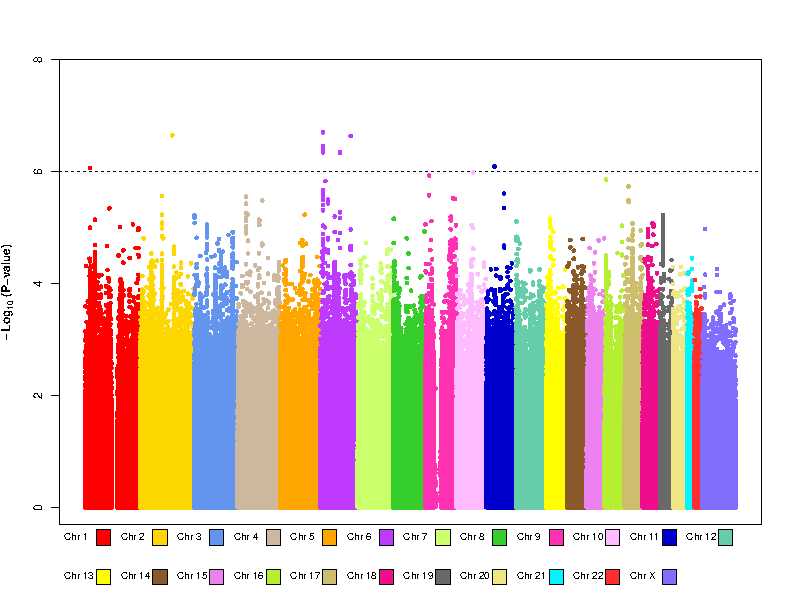

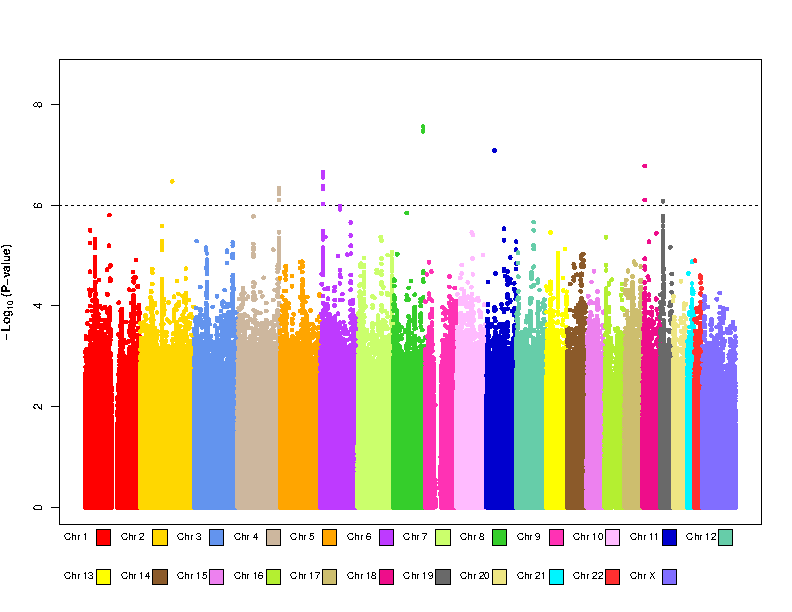

**d**

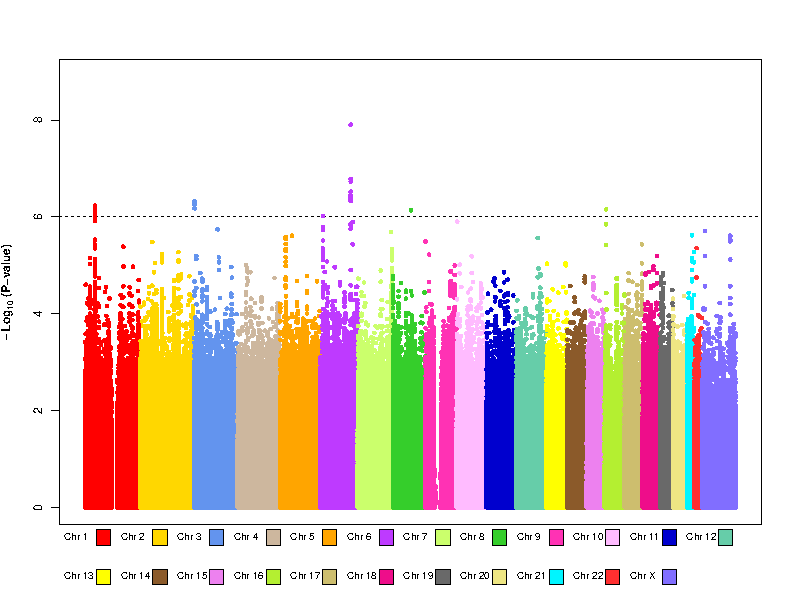

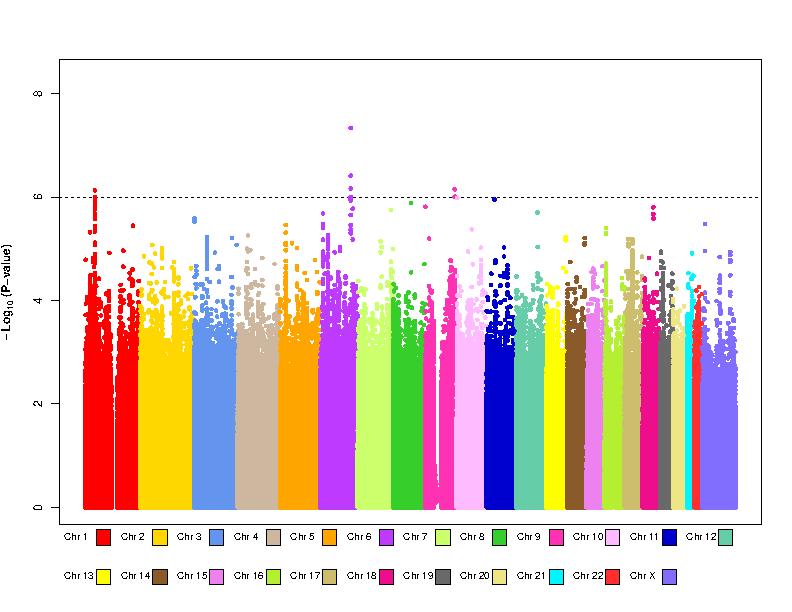

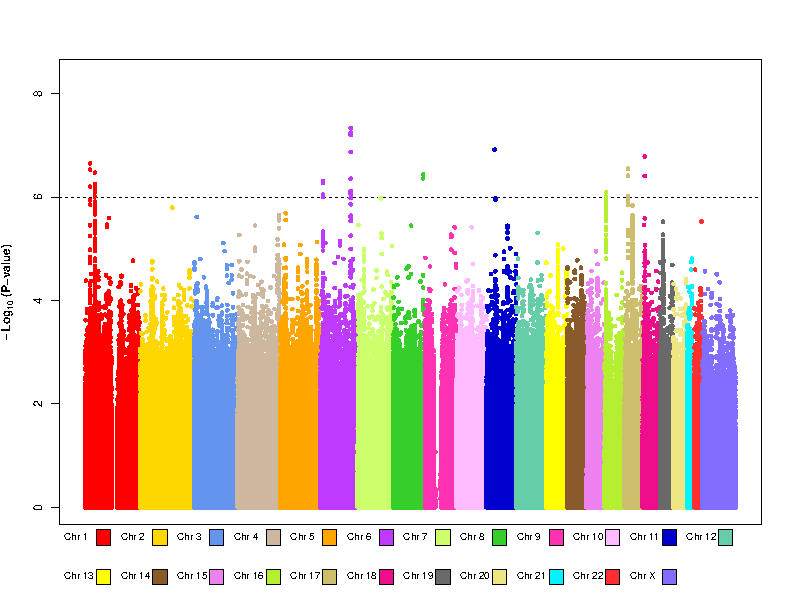

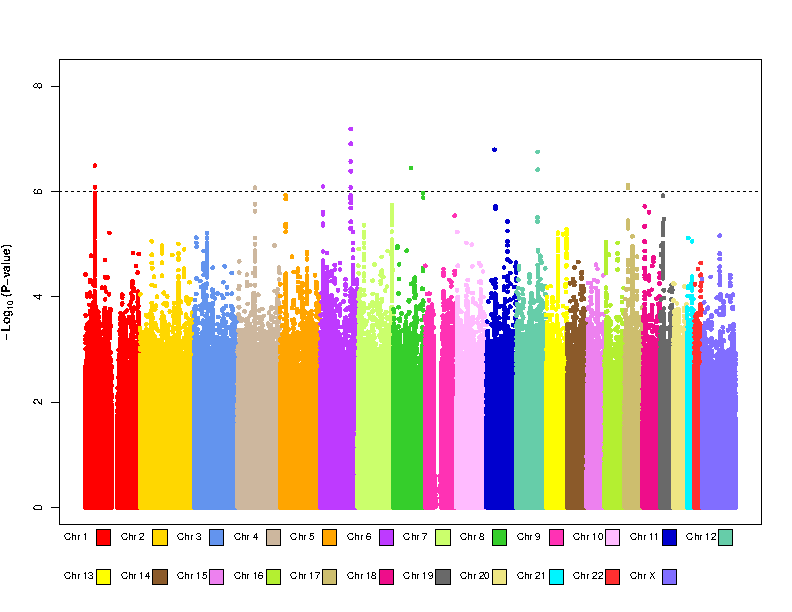

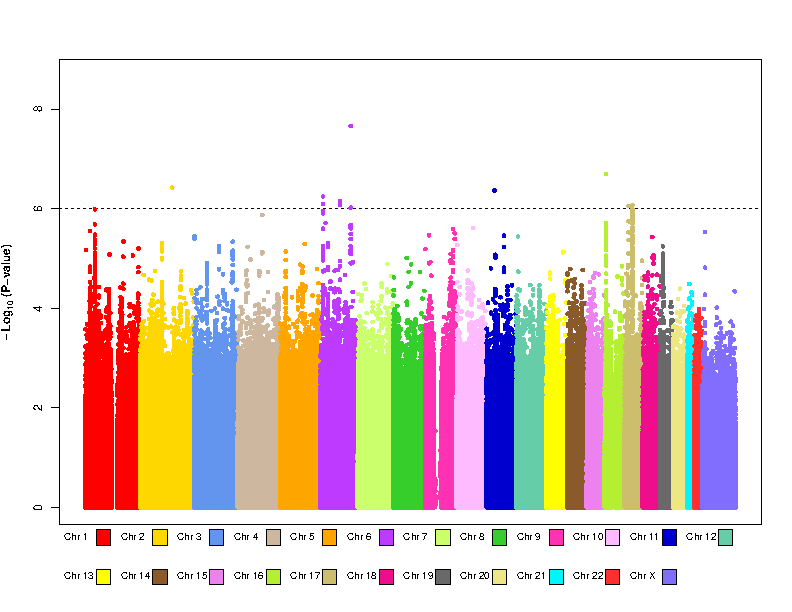

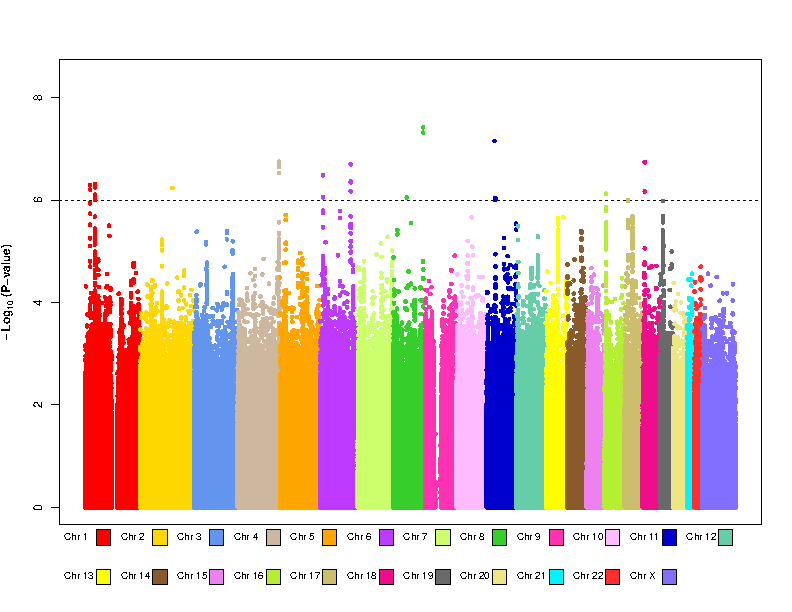

**e**

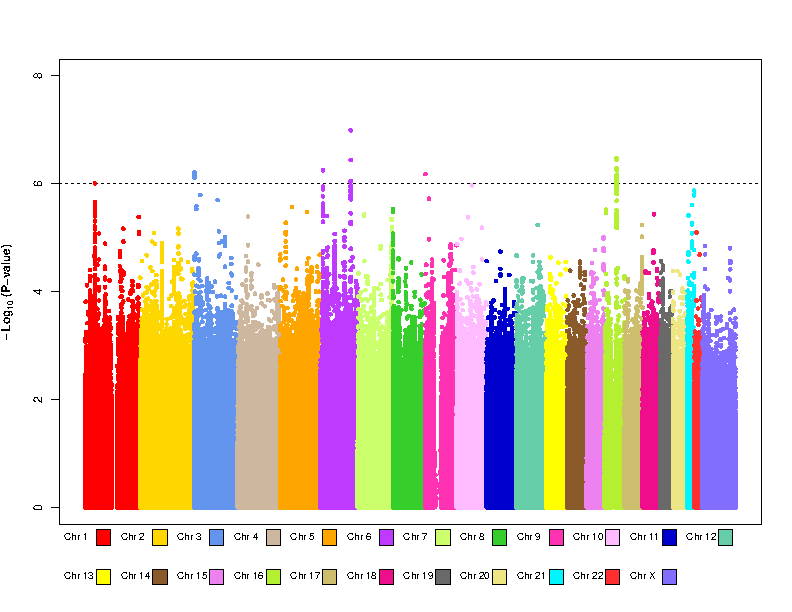

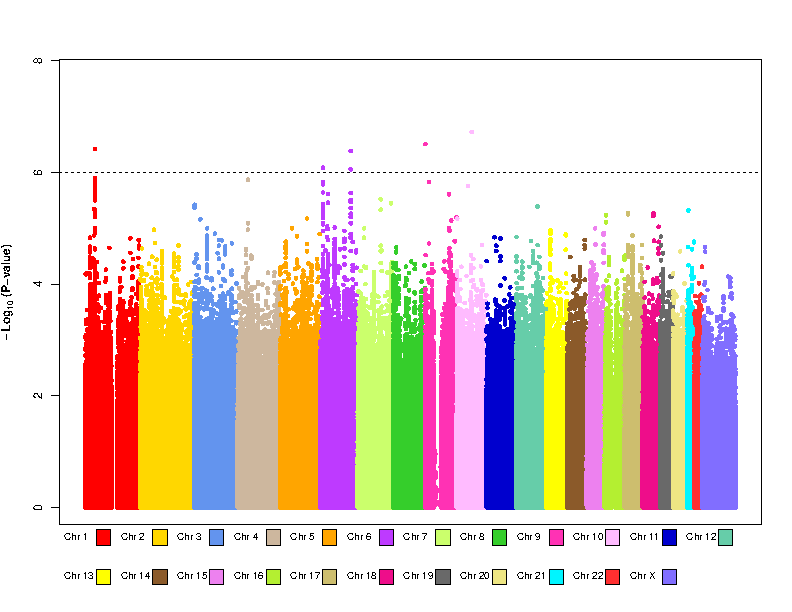

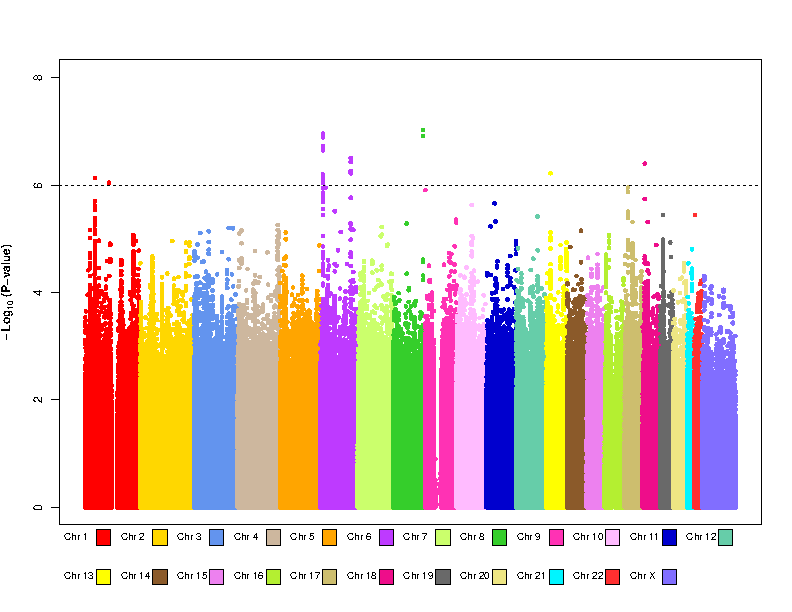

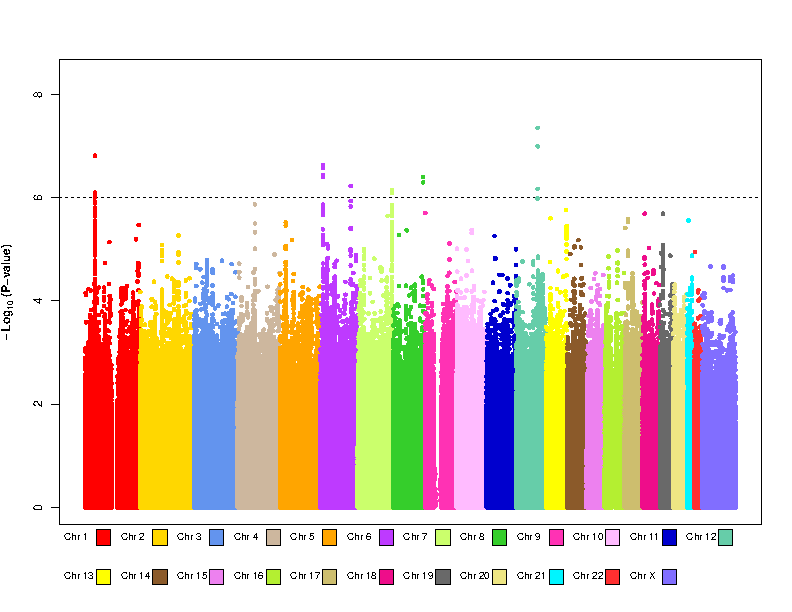

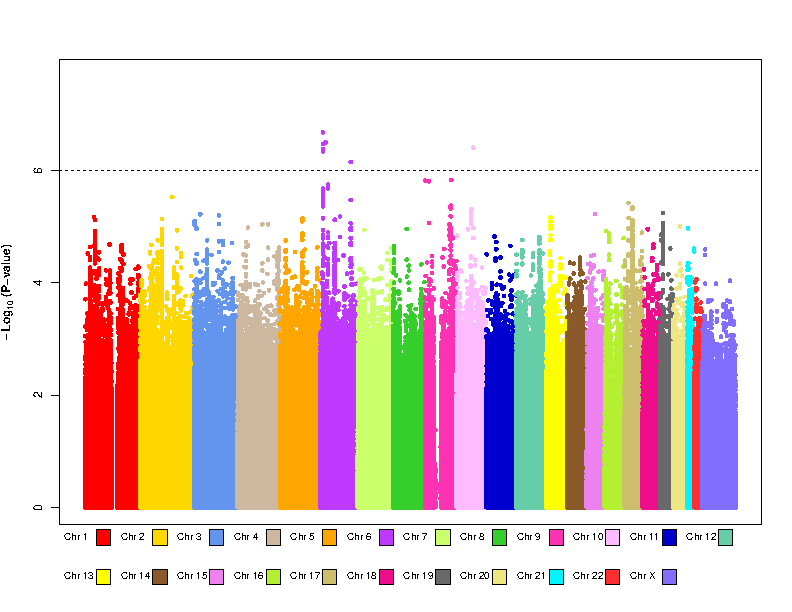

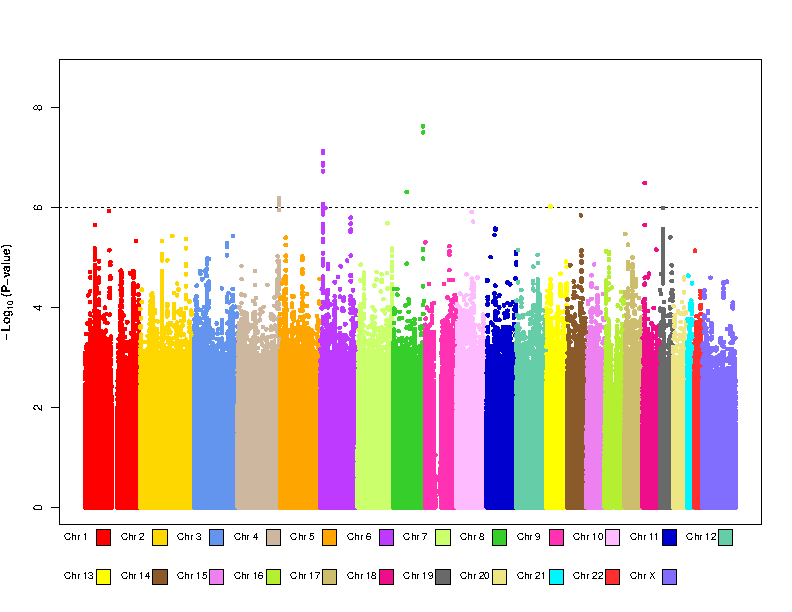

Manhattan plots of the GWAS meta-analysis results. **a** = V65L; **b** = V75L; **c** = V65H; **d** = V75H; **e** = VSUM. On each panel, the plots corresponding to Model 0 to Model 5 from left to right.

**Figure S3.** Quantile-quantile plots of the *P* value of heterogeneity

**a**

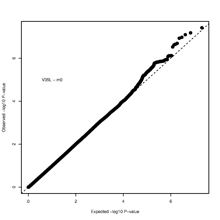

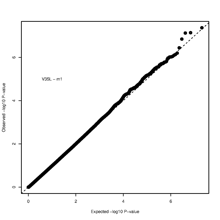

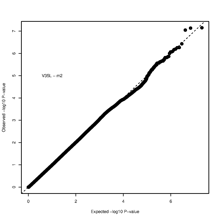

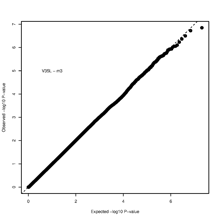

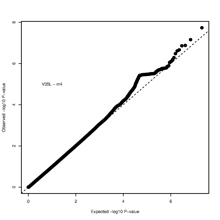

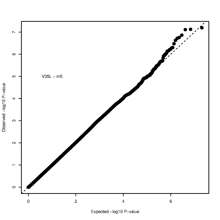

**b**

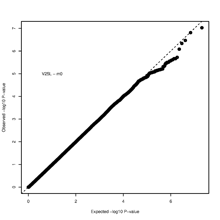

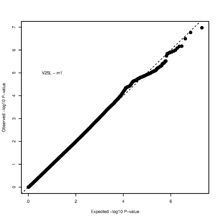

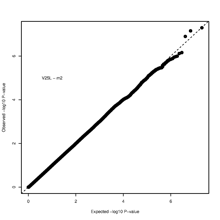

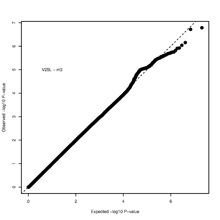

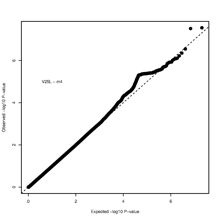

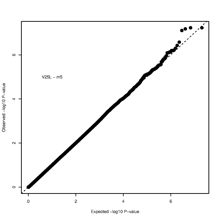

**c**

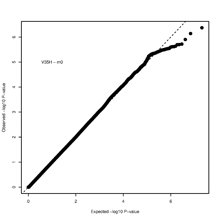

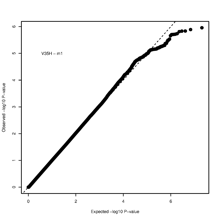

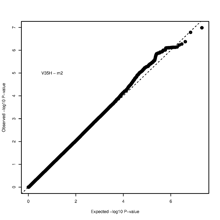

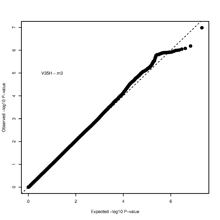

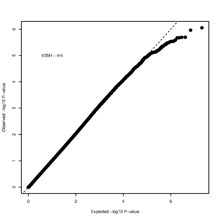

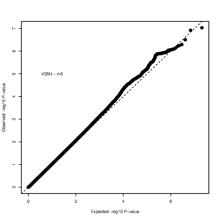

**d**

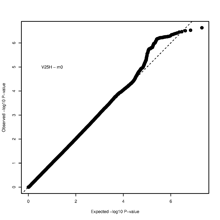

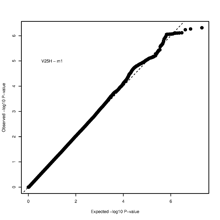

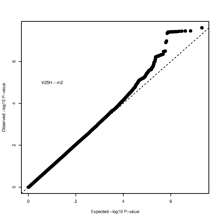

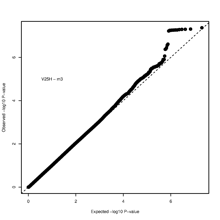

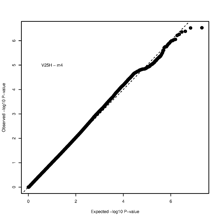

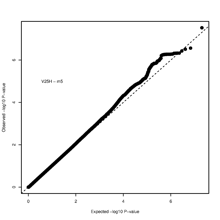

Quantile-quantile plots of the *P* value of heterogeneity. **a** = V65L; **b** = V75L; **c** = V65H; **d** = V75H. On each panel, the plots corresponding to Model 0 to Model 5 from left to right.

**References**

1.	Chen H, Fan S, Stone J, Thompson DJ, Douglas J, Li S, Scott C, Bolla MK, Wang Q, Dennis J *et al*: **Genome-wide and transcriptome-wide association studies of mammographic density phenotypes reveal novel loci**. *Breast Cancer Res* 2022, **24**(1):27.

2.	Michailidou K, Lindstrom S, Dennis J, Beesley J, Hui S, Kar S, Lemacon A, Soucy P, Glubb D, Rostamianfar A *et al*: **Association analysis identifies 65 new breast cancer risk loci**. *Nature* 2017, **551**(7678):92-94.

3.	Pulit SL, Stoneman C, Morris AP, Wood AR, Glastonbury CA, Tyrrell J, Yengo L, Ferreira T, Marouli E, Ji Y *et al*: **Meta-analysis of genome-wide association studies for body fat distribution in 694 649 individuals of European ancestry**. *Hum Mol Genet* 2019, **28**(1):166-174.

4.	Warner ET, Rice MS, Zeleznik OA, Fowler EE, Murthy D, Vachon CM, Bertrand KA, Rosner BA, Heine J, Tamimi RM: **Automated percent mammographic density, mammographic texture variation, and risk of breast cancer: a nested case-control study**. *NPJ Breast Cancer* 2021, **7**(1):68.

5.	Day FR, Thompson DJ, Helgason H, Chasman DI, Finucane H, Sulem P, Ruth KS, Whalen S, Sarkar AK, Albrecht E *et al*: **Genomic analyses identify hundreds of variants associated with age at menarche and support a role for puberty timing in cancer risk**. *Nat Genet* 2017, **49**(6):834-841.

6.	Day FR, Ruth KS, Thompson DJ, Lunetta KL, Pervjakova N, Chasman DI, Stolk L, Finucane HK, Sulem P, Bulik-Sullivan B *et al*: **Large-scale genomic analyses link reproductive aging to hypothalamic signaling, breast cancer susceptibility and BRCA1-mediated DNA repair**. *Nat Genet* 2015, **47**(11):1294-1303.

7.	Sieh W, Rothstein JH, Klein RJ, Alexeeff SE, Sakoda LC, Jorgenson E, McBride RB, Graff RE, McGuire V, Achacoso N *et al*: **Identification of 31 loci for mammographic density phenotypes and their associations with breast cancer risk**. *Nat Commun* 2020, **11**(1):5116.

8.	Zhang H, Ahearn TU, Lecarpentier J, Barnes D, Beesley J, Qi G, Jiang X, O'Mara TA, Zhao N, Bolla MK *et al*: **Genome-wide association study identifies 32 novel breast cancer susceptibility loci from overall and subtype-specific analyses**. *Nat Genet* 2020, **52**(6):572-581.
